# Women exposure to Di(2-ethylhexyl)phthalate (DEHP) and bisphenol A (BPA) from different residing areas in Italy: data from the LIFE PERSUADED project

**DOI:** 10.1101/2022.05.31.22275004

**Authors:** Fabrizia Carli, Sabrina Tait, Luca Busani, Demetrio Ciociaro, Veronica Della Latta, Anna Paola Pala, Annalisa Deodati, Andrea Raffaelli, Filippo Pratesi, Raffaele Conte, Francesca Maranghi, Roberta Tassinari, Enrica Fabbrizi, Giacomo Toffol, Stefano Cianfarani, Cinzia La Rocca, Amalia Gastaldelli, LIFE PERSUADED Project Group

## Abstract

Phthalates and bisphenol A (BPA) are plasticizers used in many industrial products that can act as endocrine disruptors. In the frame of the LIFE PERSUADED project, we measured the urinary concentrations of BPA and Di (2-ethylhexyl) phthalate (DEHP) metabolites in 900 Italian women enrolled between 2015 and 2017 and living in North, Centre and South of Italy in both rural and urban areas.

The whole cohort, representative of the Italian female adult population, was exposed to DEHP and BPA with measurable levels above the limit of detection (LOD) in more than 99% and 95% of the samples, respectively. The exposure patterns differed for the two chemicals in the three macro-areas with the highest urinary levels for DEHP in South compared to Central and Northern Italy and for BPA in Northern compared to Central and Southern Italy. BPA levels were higher in women living in urban areas, whereas no difference between areas was observed for DEHP. The estimated daily intake of BPA was 0.11 μg/kg per day, about 36-fold below the current temporary Tolerable Daily Intake of 4 μg/kg per day established by the European Food Safety Authority in 2015. The analysis of cumulative exposure showed a positive correlation between DEHP and BPA.

These results suggest to further limit the exposure to DEHP and BPA through specific legislative measures.

## 1. Introduction

Phthalates and bisphenols have been manufactured in large quantities by industry since the early 1900s as additives to make plastic polymers variably rigid, transparent, elastic, soft or resistant (Chou and Wright, 2006; Guart et al., 2011). For these properties phthalates and bisphenols, of which di 2-ethylhexyl-phthalate (DEHP) and bisphenol A (BPA) are the most common, have a wide use and are found in many industrial products such as toys, medical devices and plastic bottles, but also in personal care products, cosmetics, perfumes and food packaging (Halden, 2010).

Food contact materials represent the 20% of total exposure to BPA, with canned food as the main source of external dietary exposure and thermal paper the second (EFSA Panel on Food Contact Materials, 2015). In 2016 the Committee for Risk Assessment (RAC) of the European Chemicals Agency published a new regulation (Commission Regulation 2016/2235 of 12 December 2016) regarding the Registration, Evaluation, Authorisation and Restriction of Chemicals (REACH) which stated that BPA “shall not be placed on the market in thermal paper in a concentration equal to or greater than 0,02% by weight after 2 January 2020”. In 2018 following the opinion published by EFSA, the specific migration limit of BPA for plastic materials and articles was decreased from 0.6 mg/kg of food to 0.05 mg/kg of food (Commission Regulation(EU) 2018/213 of 12 February 2018).

With regard to DEHP, the European Food Safety Authority (EFSA) in 2005 and then in 2019 reported that food is the main source of exposure (EFSA Panel on Food Contact Materials et al., 2019).

DEHP and BPA were included in the list of substances subjected to authorization under the REACH regulation and candidates for substitution as of very high concern (ECHA/PR/18/01) (ECHA/ED/108/2014) and both are recognized as endocrine-disrupting chemicals (EDCs) with effects on the endocrine and reproductive systems, thyroid, liver, kidneys, nervous system and cancer development (Kumar et al., 2020; Lin et al., 2011; Lobstein and Brownell, 2021; Mengozzi et al., 2019; Yilmaz et al., 2020). Further, both DEHP and BPA are considered “obesogenic” EDCs due to the associations of their exposure with metabolic syndrome, obesity, type 2 diabetes mellitus and hepatic fat accumulation (Foulds et al., 2017; Heindel et al., 2015). Due to the recent evidence on adverse effects of BPA on the immunological system, the European Food Safety Authority is re-evaluating the Tolerable Daily intake suggesting to lower the value from the current 4 μg/kg day to 0.04 ng/Kg day (EFSA Panel on Food Contact Materials, 2021).

DEHP and BPA are rapidly metabolized after ingestion. DEHP is hydrolysed to mono(2-ethylhexyl) phthalate (MEHP) by unspecific lipases (Frederiksen et al., 2007) then MEHP is oxidized to mono(2-ethyl-5-hydroxyhexyl) phthalate (MEHHP) and to mono(2-ethyl-5-oxohexyl) phthalate (MEOHP). Monoesters and the oxidative metabolites are conjugated to form hydrophilic glucuronide conjugate metabolites, which are easily excreted in urine (Duty et al., 2005; Frederiksen et al., 2007; Koch et al., 2004; Wittassek and Angerer, 2008). The major metabolic pathway of BPA in humans is the BPA-glucuronidation by the enzyme UDP-glucuronyl; BPA can also be conjugated via sulfation by sulfotransferases (Ho et al., 2017). In addition, in vivo and in vitro studies suggest that BPA may be subjected to oxidation with the production of metabolites among which 4-methyl-2,4-bis(p-hydroxyphenyl)pent-1-ene (MBP) was found to increase estrogenic activity (Yoshihara et al., 2004) as well as to trigger endoplasmic reticulum (ER) stress (Huang et al., 2021).

Biomonitoring (HBM) is a powerful investigative tool allowing to evaluate human health risk to chemical exposure through the measurement of either the chemical substances, their metabolites or markers of subsequent health effects in body fluids or tissues (Choi et al., 2015). Humans are exposed to both BPA and

DEHP, however, the real exposure in Italy is still unknown because of the limited number of studies performed up to now, or the limited number of subjects enrolled in the studies. The LIFE PERSUADED project is a large HBM study with the aim to establish the exposure to phthalates and BPA in the Italian population throughout the territory and provide background levels in mothers and their children in three different geographical macro-areas. To accomplish this, 900 mother-child pairs were enrolled from 2015 to 2017 from urban and rural areas located in the North, Centre and South of Italy, (La Rocca et al., 2018). We measured BPA and DEHP levels (by the quantification of its metabolite concentrations, MEHP, MEOHP and MEHHP), in urine samples by high sensitivity mass spectrometry.

In this paper, we report the HBM results for the mothers with comparison of exposure levels according to living areas, which complete the data already published for the children (Tait et al., 2020; Tait et al., 2021). Results of this large HBM study may support the adoption of legislative measures aimed at limiting the exposure to DEHP and BPA.

## 2. Materials and Methods

### 2.1 Study population

In the LIFE PERSUADED project, a HBM study was performed enrolling 900 Italian healthy mother-child pairs between 2015 and 2017 equally distributed in Northern, Central or Southern Italy (La Rocca et al., 2018).

Subjects were also selected for equally residing in urban or rural areas, as defined by population density (> or < 150 inhabitants /Km2) and number of inhabitants (> or < 50000 inhabitants) on the basis of the Italian Institute of Statistics (ISTAT) database. Included subjects had to be at their residence for at least 6 months prior the enrolment. Mothers were also equally distributed according to their children sex and age category (4-6 yrs, 7-10 yrs, 11-14 yrs). The enrolment was carried out in different Italian regions by family pediatricians of the Italian Health System, who voluntarily joined the HBM study through the *Associazione Culturale Pediatri* (ACP, http://www.acp.it) and the Federazione Italiana Medici Pediatri Marche (FIMPM, http://www.fimpmarche.it) networks as previously described (La Rocca et al., 2018).

Inclusion criteria were healthy status, being mother of children between 4 and 14 years and with a body mass index between 5^th^ and 85^th^ percentile. Mothers in gestational status or in breastfeeding were excluded.

The whole study was approved by the Ethical Committee of the Istituto Superiore di Sanità. The enrolled mothers signed an informed consent to provide a first-morning urine sample for the measurement of DEHP and BPA exposure and filled a questionnaire with information on residential environment, socio-demographic and lifestyle data as well as on food habits. Participants were assigned an alphanumeric code to guarantee anonymity (La Rocca et al., 2018).

Subjects were excluded from final analysis if urine sample was not available. Final analysis was performed on 898 mothers equally distributed in North (N=300: n=150 rural and n=150 urban), Centre (N=299: n=149 rural and n=150 urban) and South (N=299: n=149 rural and n=150 urban) areas. BMI data were available for 879 women, whereas only 655 women declared their age in the questionnaires.

### 2.2 Sample Collection and Measurements

Concentration of DEHP metabolites and BPA was measured in first-morning urine samples that each pediatrician stored at -20 °C and then shipped in dry ice in batches to the CNR in Pisa for centralized chemical analysis. First morning urine sampling has been chosen in agreement with guidelines for HBM studies (Becker et al., 2014).

Upon arrival to central laboratory, samples were thawed overnight at 4°C and then aliquoted in polypropylene (PP) tubes (BPA- and phthalates free) for creatinine and mass analytical determination (stored at –20 °C until analysis). Two additional tubes (1.8 ml each) were stored in the bio-bank of the Institute of Clinical Physiology of CNR in Pisa (https://www.ifc.cnr.it/index.php/it/le-aree-di-competenza/la-biomedicina/bio-banca).

Creatinine concentrations in urine samples were determined by the Jaffe’s method through an AU400 Beckman instrument (Beckman, Brea, CA, USA) and were all in the range 0.3-3 g/L (WHO, 1996).

For both DEHP metabolites and BPA analysis, 500 µl of urines were deconjugated by adding 2 µl β-glucuronidase (from Helix Pomatia enzyme aqueous solution, ≥ 100.000 units/mL; Sigma Aldrich, St Louis, MO, USA) and 375 μl of ammonium acetate buffer (NH4CH3COO−) 1 M (Sigma Aldrich), at pH=5, and incubating at 37°C overnight. Internal standards (^13^C-DEHP metabolites and D_16_-BPA) were added in the samples that were extracted and purified by C18-SPE column (SPE cartridge C18 ODS 3 ml tubes 200 mg, Agilent, Santa Clara, CA), eluting with acetonitrile and methanol and dried under a gentle nitrogen flux (Carli et al., 2022).

#### 2.2.1 Determination of phthalate levels

DEHP metabolites (e.g. MEHP, MEHHP and MEOHP) were determined following the analytical method described in detail in (Carli et al., 2022). Briefly, after reconstitution with the acetonitrile:water solution (1:9, v:v), samples were injected in a UHPLC instrument (UHPLC 1290 infinity coupled with Quadrupole Time-of-Flight QTOF 6540, Agilent, Santa Clara CA, USA) equipped with Agilent ZORBAX SB-Phenyl 2.1×100mm 1.8-Micron.

Limit of Detection and Quantification (LOD and LOQ) were determined according to IUPAC guidance and US Environmental Protection Agency (https://www.epa.gov/cwa-methods/procedures-detection-and-quantitation-documents). We found that LOD and LOQ were for MEHP 0.28 and 0.58 ng/ml, for MEOHP 0.18 and 0.48 ng/ml and for MEHHP 0.11 and 0.24 ng/ml, respectively (Carli et al., 2022).

Information on background levels, blank and spike recoveries, calibration curves and quality control samples are reported in (Carli et al., 2022).

#### 2.2.2 Determination of BPA levels

After LC-MS analysis, samples were dried under a gentle nitrogen flux and then derivatizated with 10 µl of BSTFA 1%TMCS and 50 µl of acetonitrile (Sigma Aldrich, Merck KGaA, Darmstadt, Germany (Carli et al., 2022). BPA analysis was performed by GCMS (7890A coupled with 5975 Agilent, Santa Clara CA) equipped with a capillary column (DB-5MS J&W, l 30 m; i.d. 0.25 mm; film thickness 0.25 μm) in selected ion monitoring (SIM) mode by analyzing the fragment ions 357 for BPA and 368 for D16-BPA (CDN isotopes, Pointe-Claire, Quebec CDN).

The Limit of Detection (LOD) and the Limit of Quantification (LOQ) were calculated according to U.S. EPA procedure (U.S. EPA, 2017) and were 0.157 ng/ml and 0.523 ng/ml, respectively (Carli et al., 2022).

Both analytical methods used to determine DEHP metabolites and BPA were validated in a proficiency test (ICI/EQUAS) in the frame of the HBM4EU Project (https://www.hbm4eu.eu/online-library/), as previously reported (Carli et al., 2022).

### 2.3 Relative metabolic rates and percentage fractions

Calculation of relative metabolic rates (RMR) and percentage fractions of DEHP metabolites was previously described (Tait et al., 2020). Briefly, to derive RMRs, each DEHP metabolite concentration was converted in molar concentration and conversion rates were calculated according to the two-step metabolic transformations:

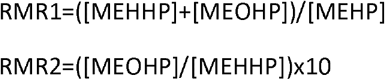

Percentage fractions of each DEHP metabolite were calculated with respect to the sum of all metabolites.

### 2.4 Derivation of Reference values

Reference values (RV95) for both DEHP as sum of its metabolites and BPA were calculated as 95^th^ percentile with corresponding 95% confidence intervals, as previously described (Tait et al., 2020). RV95 was calculated for total population, macro-areas, areas and age class.

### 2.5 Daily intake of BPA

Total daily intake (TDI) of BPA was calculated by using the EFSA equation based on BPA concentration in urine volume (EFSA Panel on Food Contact Materials, 2015):

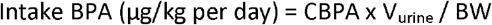

where CBPA is the BPA concentration in urine sample (µg/L), V_urine_ is the daily urinary volume [(1.2 (L) x BW (kg)]. Among the 898 eligible enrolled women, 19 did not provide their BW, thus the calculation was performed on 879 women.

### 2.6 Statistical analysis

Statistical analysis was performed with STATA 14.2 (StataCorp, 4905 Lakeway Drive, College 17Station, Texas, TX, USA) setting significance at p < 0.05 for all the statistical tests performed.

Since data are not normally distributed, non-parametric tests were performed to evaluate statistical differences among different groups. For two-levels strata (e.g., area), the Mann-Whitney test was performed, whereas for three-levels strata (e.g., macro-area), the Kruskal-Wallis test was performed with Dunn’s post hoc evaluation where applicable. Differences between groups were calculated for both unadjusted and creatinine-adjusted urinary levels.

Correlation analysis was performed by Spearman’s test with Bonferroni correction.

For additional analyses, age and BMI were categorized as follows: age (20-30, 30-40, 40-50, >50), BMI (<25, 25-30, >30).

Analytical data on DEHP metabolites and BPA in the Italian women involved in this study will be made available on the Information Platform for Chemical Monitoring of the European Commission (https://ipchem.jrc.ec.europa.eu) as already done for DEHP data on children.

## 3. Results

### 3.1 Characteristics of the cohort

The clinical characteristics of the mothers participating to the study are shown in **Table 1**. The median age was 41 years ranging from 24 to 67 years. Mothers from Southern Italy were younger compared to those living in Central Italy and mothers living in urban areas were older than those in rural areas. Although the great majority of these women was lean (median BMI of 22.27 kg/m^2^; 20.34-24.80 interquartile range), BMI data showed a significant trend to higher values from North to South of Italy, while no significant difference was observed between mothers residing in rural or urban areas.

**Table 1.** Distribution of enrolled women age and BMI in Italian macro-areas and areas. Median values and interquartile range are indicated; p-values for significant differences among groups are indicated in the beside column.

|  | AGE |  |  | BMI |  |  |
| --- | --- | --- | --- | --- | --- | --- |
|  | N | P50 (P25-P75) | p-value | N | P50 (P25-P75) | p-value |
| **TOT** | 655 | 41 (38-44) |  | 879 | 22.27 (20.34-24.80) |  |
| **North** | 254 | 41 (38-44) |  | 295 | 21.83 (20.02-24.11)<sup>a</sup> | 0.0062<sup>a vs c</sup> |
| **Centre** | 193 | 41 (38-44)<sup>a</sup> |  | 290 | 22.46 (20.20-24.61)<sup>b</sup> | 0.0348<sup>b vs a</sup> |
| **South** | 208 | 40 (36-44)<sup>b</sup> | 0.0390<sup>b vs a</sup> | 293 | 22.77 (21.09-25.95)<sup>c</sup> | <0.0001<sup>c vs a</sup> |
| **Rural** | 319 | 40 (36-43)<sup>a</sup> |  | 439 | 22.27 (20.57-25.00) |  |
| **Urban** | 336 | 41 (38-45)<sup>b</sup> | 0.0024<sup>b vs a</sup> | 440 | 22.27 (20.31-24.70) |  |

<sup>a,b,c</sup> Different superscript letters indicate statistically significant differences between groups; superscript letters beside the p-values indicate the corresponding pairwise comparison.

### 3.2 Exposure to phthalates

#### 3.2.1 Phthalates levels in Italian women

In this cohort of Italian women, DEHP was detected in over 99% of the subjects (**Table 2**): MEHP values above LOD were detected in about 99% of the population and the secondary metabolites MEHHP and MEOHP in about 97% and 99% of the population, respectively. **Table 2** reports unadjusted urine levels in μg/L, and levels adjusted to creatinine content, in μg/g, for each DEHP metabolite and their sum. As expected, the secondary DEHP metabolite MEHHP was found at higher levels compared to the other metabolites.

**Table 2.** DEHP metabolite levels in urine samples of 898 Italian women. In the table are reported the percentage of the samples above the Limit of Detection (LOD), the geometric mean (GM) with the 95% confidence interval (CI), the median (P50) with the interquartile range (P25-P75). For each metabolite, the first row refers to the unadjusted concentration in urine (μg/L) and the second row to the creatinine-adjusted concentration (μg/g).

| METABOLITE | UNIT | >LOD(%) | GM (95% CI) | P50 (P25-P75) |
| --- | --- | --- | --- | --- |
| MEHP | µg/L | 99.33% | 4.64 (4.39-4.91) | 5.10 (3.37-7.41) |
|  | µg/g crea | 99.33% | 4.30 (4.04-4.58) | 4.50 (2.92-6.84) |
| MEHHP | µg/L | 97.44% | 18.02 (16.96-19.14) | 17.97 (10.49-31.69) |
|  | µg/g crea | 97.44% | 16.55 (15.70-17.45) | 16.11 (10.23-26.44) |
| MEOHP | µg/L | 98.87% | 5.53 (5.21-5.87) | 5.62 (3.20-9.64) |
|  | µg/g crea | 98.87% | 5.11 (4.84-5.38) | 4.91 (3.14-7.98) |
| Σ PHTHALATES | µg/L | 99.33% | 29.69 (28.18-31.28) | 29.47 (18.10-47.64) |
|  | µg/g crea | 99.33% | 27.52 (26.27-28.82) | 26.58 (17.64-41.18) |

#### 3.2.2 Phthalates levels by residing area

Significant differences were found among women residing in the different macro-areas of Italy. MEHP, MEHHP, MEOHP and sum of the DEHP metabolites levels were significantly higher in women living in Southern compared to those living in the Northern and Central Italy, for both unadjusted and adjusted urinary concentrations (except adjusted MEHP concentrations). **Figure 1** shows the distribution of DEHP metabolites adjusted to creatinine content (µg/g) in North, Centre and South of Italy whereas **Table 3** reports data of the sum of DEHP metabolites in each macro-area and area (single DEHP metabolites data are reported in **Supplementary Tables 1 and 2**).

**Figure 1.**
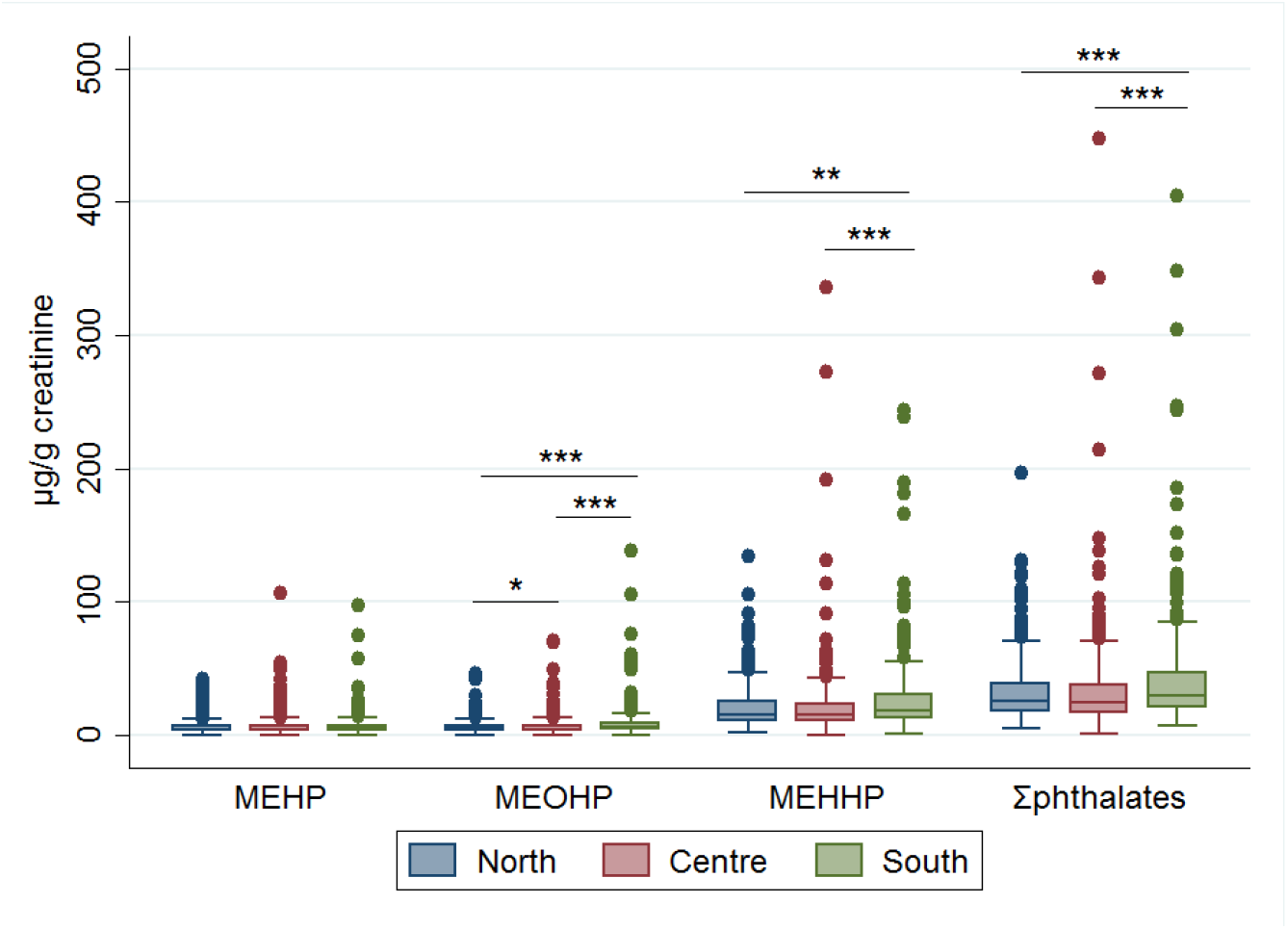
Box plots for DEHP metabolites in urine samples of Italian women residing in the North, Centre and South Italy. Data were normalized to urine creatinine concentration (µg/g). Two outlier values were excluded from the graph for a better visualization. Asterisks indicate the level of significance: *p<0.05, **p<0.01 and ***p<0.001.

**Table 3.** Levels of the sum of DEHP metabolites in urine samples of women residing in the North (N=300), Centre (N=299) or South (N=299) of Italy. In the table are reported the geometric mean (GM) with the 95% CI and the median (P50) with the interquartile (P25-P75) range. Both unadjusted (µg/L) and creatinine-adjusted concentrations (µg/g) are reported.

|  | UNIT | GM (95% CI) | P50 (P25-P75) | p-value |
| --- | --- | --- | --- | --- |
| MACRO-AREA |  |  |  |  |
| North<sup>a</sup> | µg/L | 26.72 (24.54-29.09) | 25.61 (15.80-41.79)<sup>a</sup> | <0.0001<sup>a vs b</sup> |
| Centre<sup>a</sup> |  | 27.65 (25.23-30.31) | 26.85 (17.29-45.79)<sup>a</sup> | 0.0001<sup>a vs b</sup> |
| South<sup>b</sup> |  | 35.43 (32.33-38.83) | 35.80 (21.64-56.80)<sup>b</sup> |  |
| North<sup>a</sup> | µg/g crea | 26.07 (24.22-28.06) | 25.28 (17.53-38.82)<sup>a</sup> | 0.0007<sup>a vs b</sup> |
| Centre<sup>a</sup> |  | 25.28 (23.30-27.44) | 24.39 (15.88-37.96)<sup>a</sup> | 0.0001<sup>a vs b</sup> |
| South<sup>b</sup> |  | 31.62 (29.09-34.37) | 28.91 (20.31-46.47)<sup>b</sup> |  |
| AREA |  |  |  |  |
| Rural | µg/L | 29.46 (27.39-31.69) | 29.02 (18.25-46.88) |  |
| Urban |  | 29.92 (27.76-32.24) | 29.94 (17.82-49.87) |  |
| Rural | µg/g crea | 26.55 (24.86-28.35) | 25.72 (17.50-39.36) |  |
| Urban |  | 28.53 (26.72-30.45) | 27.28 (18.13-41.71) |  |

<sup>a,b,c</sup> Different superscript letters indicate statistically significant differences between groups; superscript letters beside the p-values indicate the corresponding pairwise comparison.

No significant differences were observed between women residing in urban or rural areas for the sum of DEHP metabolites (**Table 3**) and the single metabolite levels (**Supplementary Table 2**).

Data stratified by areas in each macro-area are reported in **Table 4** for the sum of the DEHP metabolites and in **Supplementary Table 3** for the single metabolites. Significant differences between residing areas were found for MEHP (ug/L or ug/g crea) in South Italy, being higher in rural than urban areas (**Supplementary Table 3**) and for the sum of DEHP metabolites, as adjusted values, in Central Italy, being higher in urban than rural areas (**Table 4**).

**Table 4.**
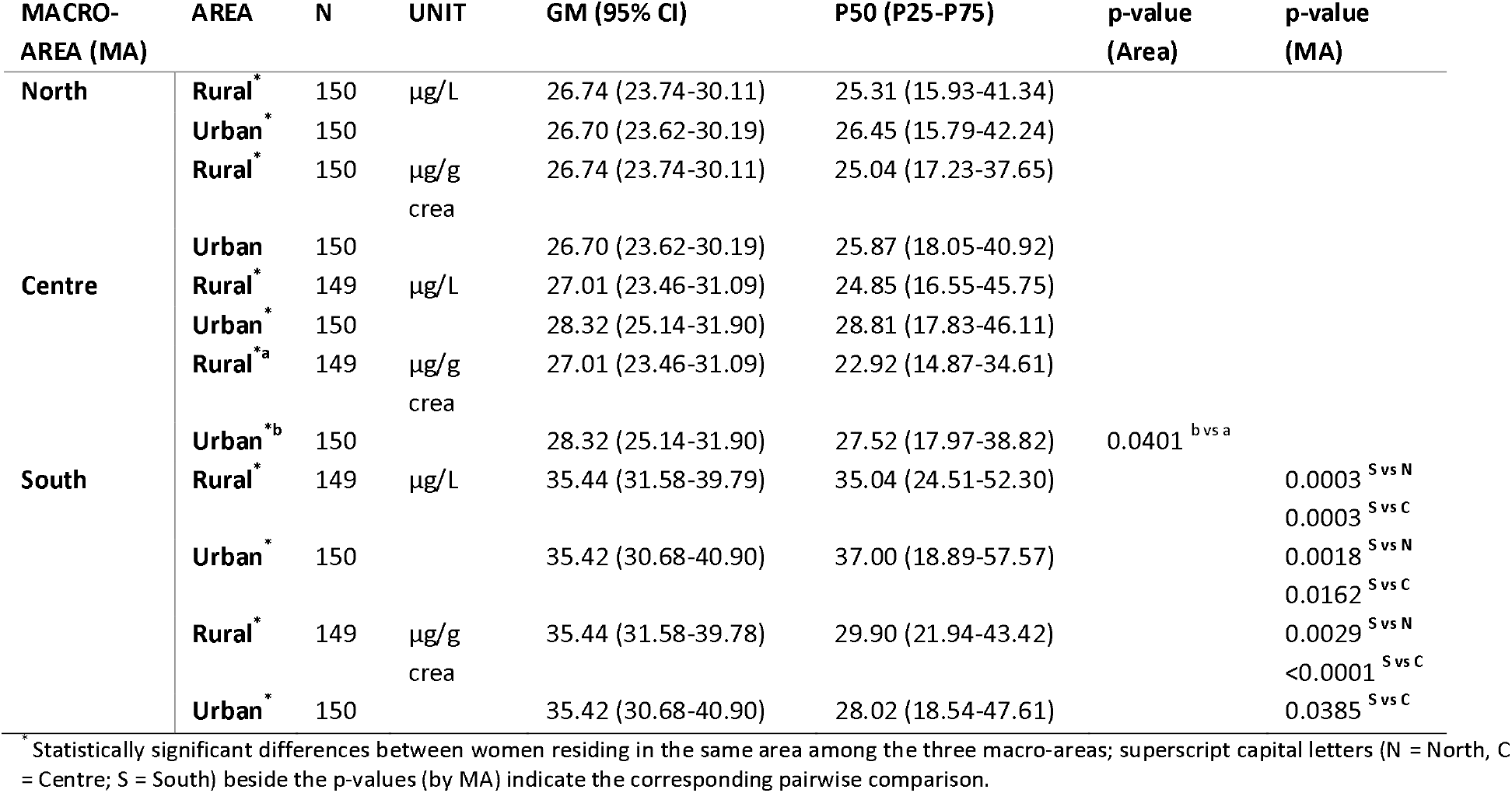
Levels of the sum of DEHP metabolites in urine samples of women residing in rural or urban Italian areas in each macro-area. In the table are reported the geometric mean (GM) values with the 95% Confidence Interval (CI), the median (P50) with the interquartile range (P25-P75). Both unadjusted (ng/ml) and creatinine-adjusted concentrations (μg/g) are reported.

For the sum of DEHP metabolites as well as for the single MEHP, MEHHP and MEOHP, higher unadjusted and creatinine-adjusted levels were observed in rural areas of Southern Italy compared to North and Centre. Further, MEOHP adjusted levels were higher in rural areas of North then of Central Italy. (**Table 4** and **Supplementary Table 3**). In urban areas of Southern Italy, women had higher unadjusted MEHHP levels compared to North and Centre and adjusted MEHHP levels compared to North; in addition, unadjusted and adjusted MEOHP levels were higher in urban areas of South compared to North and Centre (**Supplementary Table 3**), and higher unadjusted sum of DEHP metabolite levels compared to the other two areas, whereas creatinine-adjusted levels were higher only compared to Central Italy (**Table 4**).

#### 3.2.3 Phthalate metabolic rates

The analysis of the relative metabolic rates of DEHP metabolites and their molar percentages evidenced that women residing in Southern Italy had a higher metabolic conversion rate from MEHP to MEHHP+MEOHP (RMR1) compared to women from the other two macro-areas (**Table 5**). Accordingly, MEHP molar percentage in these women was significantly lower.

**Table 5.**
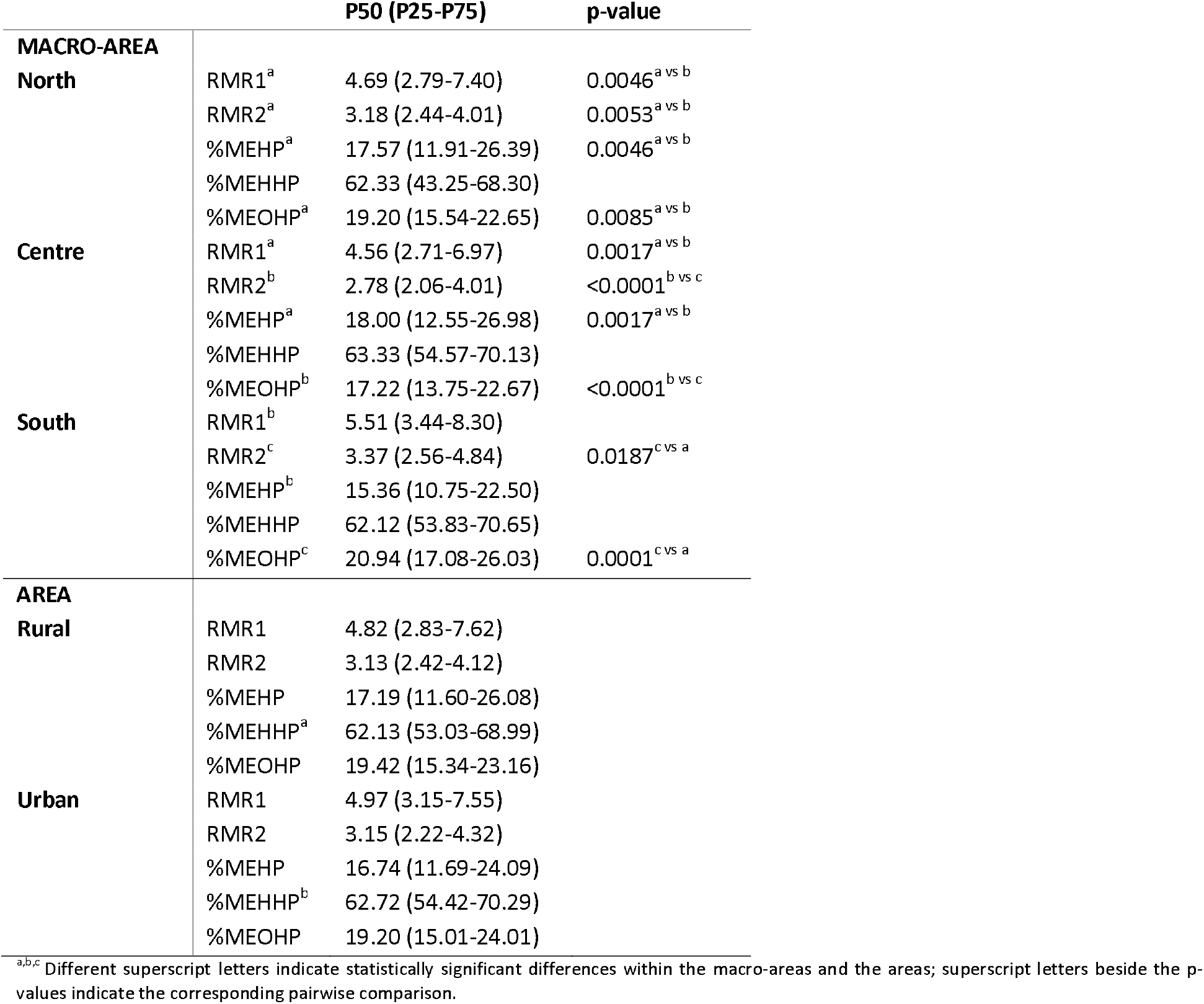
Relative metabolic rates and percentage fractions of DEHP metabolites in women residing in the three macro-areas and the two areas. Data are expressed as medians (IQ range).

The rate of the second metabolic conversion (RMR2) was significantly different among women of the three macro-areas, with increasing values from Centre to North, to South of Italy. As a consequence, the same pattern was observed for the MEOHP molar percentage. No difference in the metabolic rates were observed between women residing in rural or urban areas.

Analyzing metabolic rates and molar percentages by age categories, women aged 30-50 yrs had higher RMR1 and lower %MEHP values compared to women >50 yrs; conversely, RMR2 significantly decreased with age with higher values in women aged 20-30 yrs (**Supplementary Table 4**).

No significant difference in RMR1 and %MEHP values were observed in women according to different BMI categories. Otherwise, significantly higher RMR2 values were observed in women with a BMI > 30 compared to women with BMI < 25 (Supplementary Table 5). Accordingly, women with BMI >30 had higher % MEOHP values compared to women with BMI < 25 but also with BMI in the overweight range (25-30). Women with BMI > 30 had also lower %MEHHP values compared to overweight women.

### 3.3 Exposure to BPA

#### 3.3.1 BPA levels in Italian women

Detectable BPA levels (free and glucuronide forms) were found in 95.65% of the 898 women included in this study. Unadjusted and creatinine-adjusted concentrations are reported in **Table 6**.

**Table 6.** BPA levels in urine samples of 898 Italian women. In the table are reported the geometric mean (GM) with the 95% CI and the median (P50) with the interquartile (P25-P75) range. Both unadjusted (µg/L) and creatinine-adjusted concentrations (µg/g) are reported.

| UNITS | >LOD(%) | GM (95% CI) | P50 (P25-P75) |
| --- | --- | --- | --- |
| µg/L | 95.65% | 5.79 (5.41-6.20) | 5.76 (3.30-10.33) |
| µg/g crea | 95.65% | 5.36 (4.98-5.77) | 5.06 (3.14-9.73) |

#### 3.3.2 BPA levels by residing area

BPA levels in urine samples of women, stratified by residing macro-area and areas, are summarized in **Table 7**.

**Table 7.**
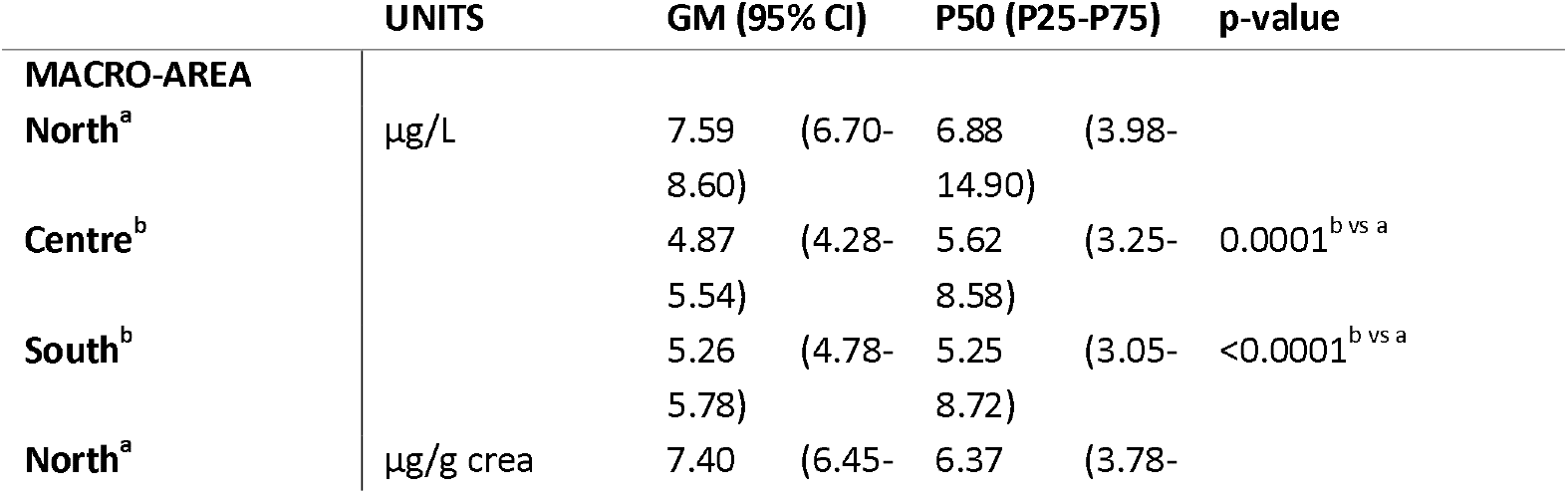
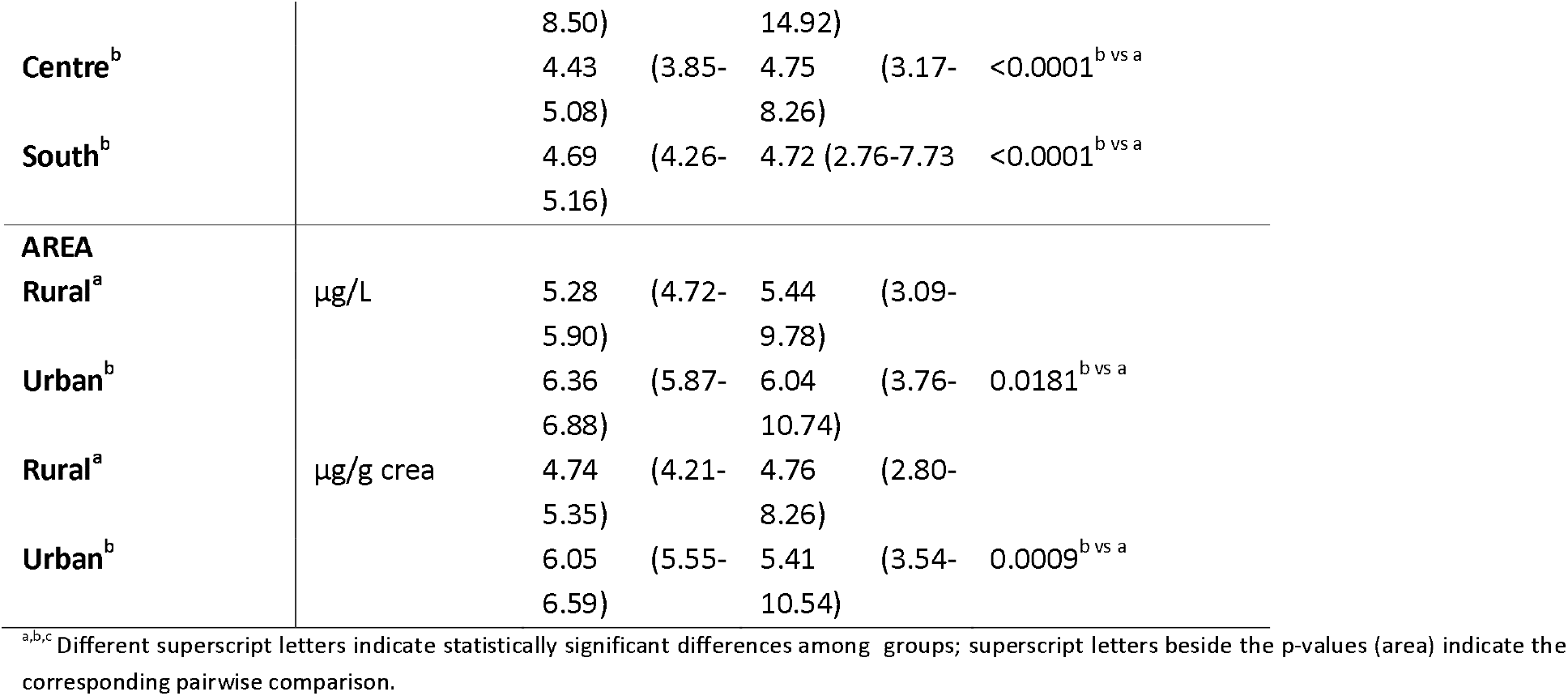
BPA levels in urine samples of women residing in the North (N=300), Centre (N=299) or South (N=299) of Italy. In the Table are reported the geometric mean (GM) with the 95% CI, the median (P50) with the interquartile range (P25-P75). Both unadjusted (µg/L) and creatinine-adjusted concentrations (µg/g) are reported.

Significantly higher BPA unadjusted and adjusted levels were found in women residing in Northern Italy compared to those residing in the other two macro-areas. Further, BPA unadjusted and adjusted levels were higher in women residing in urban than rural areas.

In Central and Southern Italy, BPA levels in women residing in urban areas were higher than in women from rural areas, both as unadjusted and adjusted values (**Figure 2** and **Table 8**). Women from rural areas of Northern Italy had higher BPA unadjusted and adjusted levels compared to women from the other two rural areas. Further, urban women from Northern Italy had higher BPA adjusted levels compared to urban women from the South of Italy.

**Figure 2.**
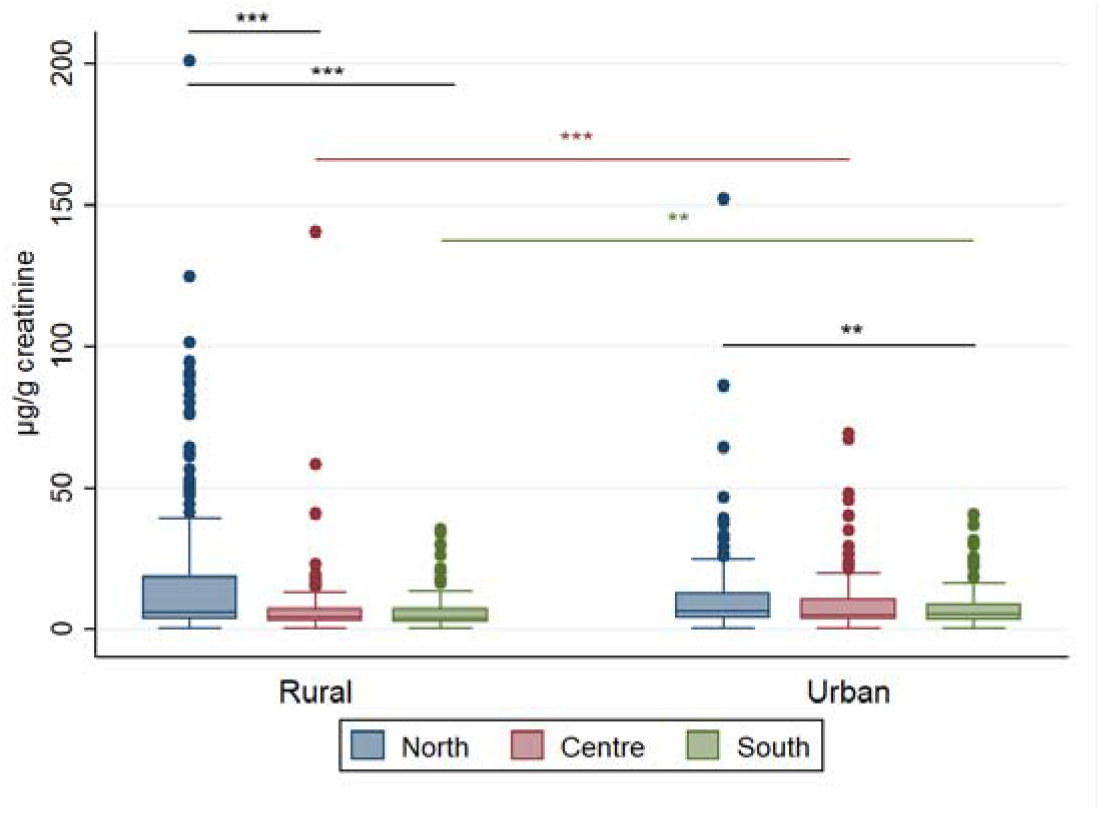
Box plots for BPA levels in urine samples of Italian women residing in the North, Centre and South Italy, in rural and urban areas. Data were normalized to urine creatinine concentration (µg/g). Two outlier values were excluded from the graph for a better visualization. Black lines and asterisks indicate statistical differences among macro-areas; red and green bars/asterisks indicate statistical differences rural and urban areas in Centre and South of Italy, respectively. Asterisks indicate the level of significance: **p<0.01 and ***p<0.001.

**Table 8.**
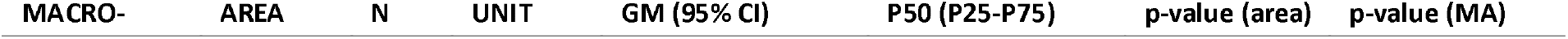
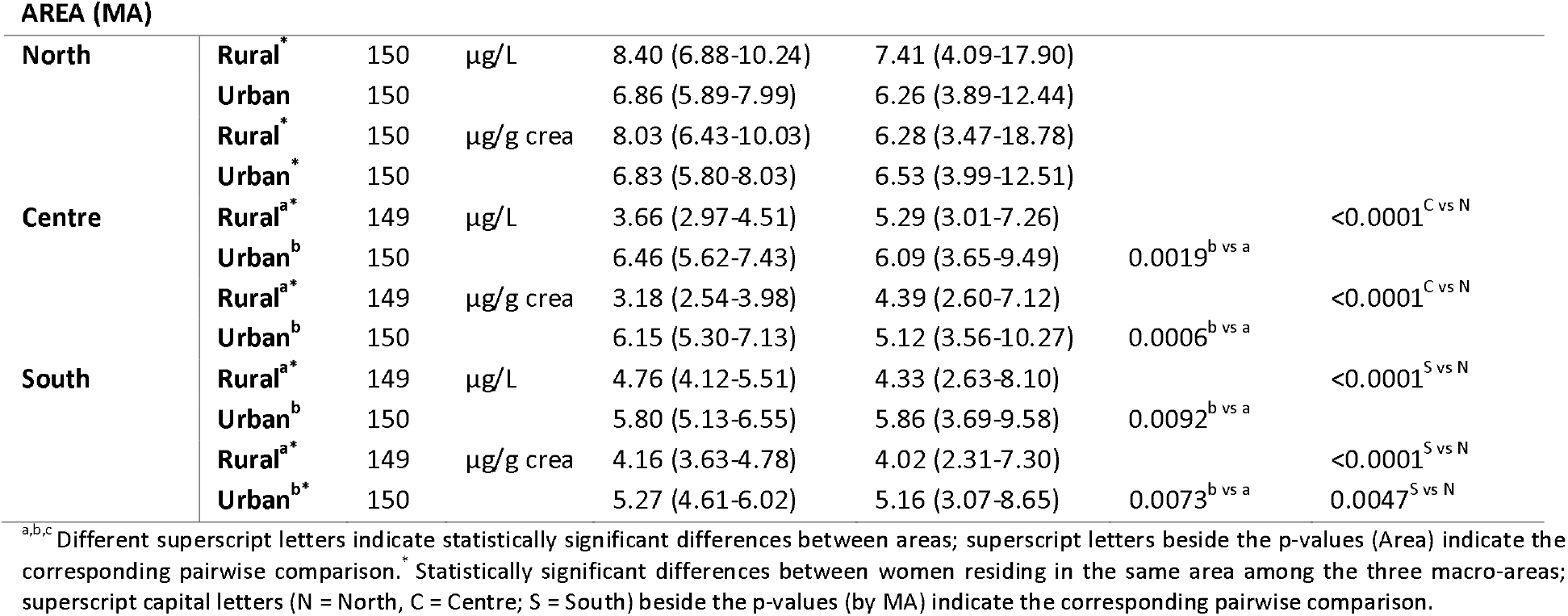
BPA levels in urine samples of women residing in rural or urban Italian areas in each macro-area. In the table are reported the geometric mean values with the 95% Confidence Interval (CI) and the median (P50) with the interquartile (P25-P75) range. Both unadjusted (ng/ml) and creatinine-adjusted concentrations (μg/g) are reported.

#### 3.3.3 Daily intake of BPA

The calculated total daily intake (TDI) for total enrolled women providing their BW (N=879) was 0.11 µg/kg bw per day as geometric mean (0.11-0.12 95% CI) (**Table 9**). Considering the current t-TDI of 4 µg/kg bw per day (EFSA Panel on Food Contact Materials, 2015), only one woman exceeded this value and 2.28% women (N=20) had a daily intake between 1 and 4 µg/kg bw per day.

**Table 9.**
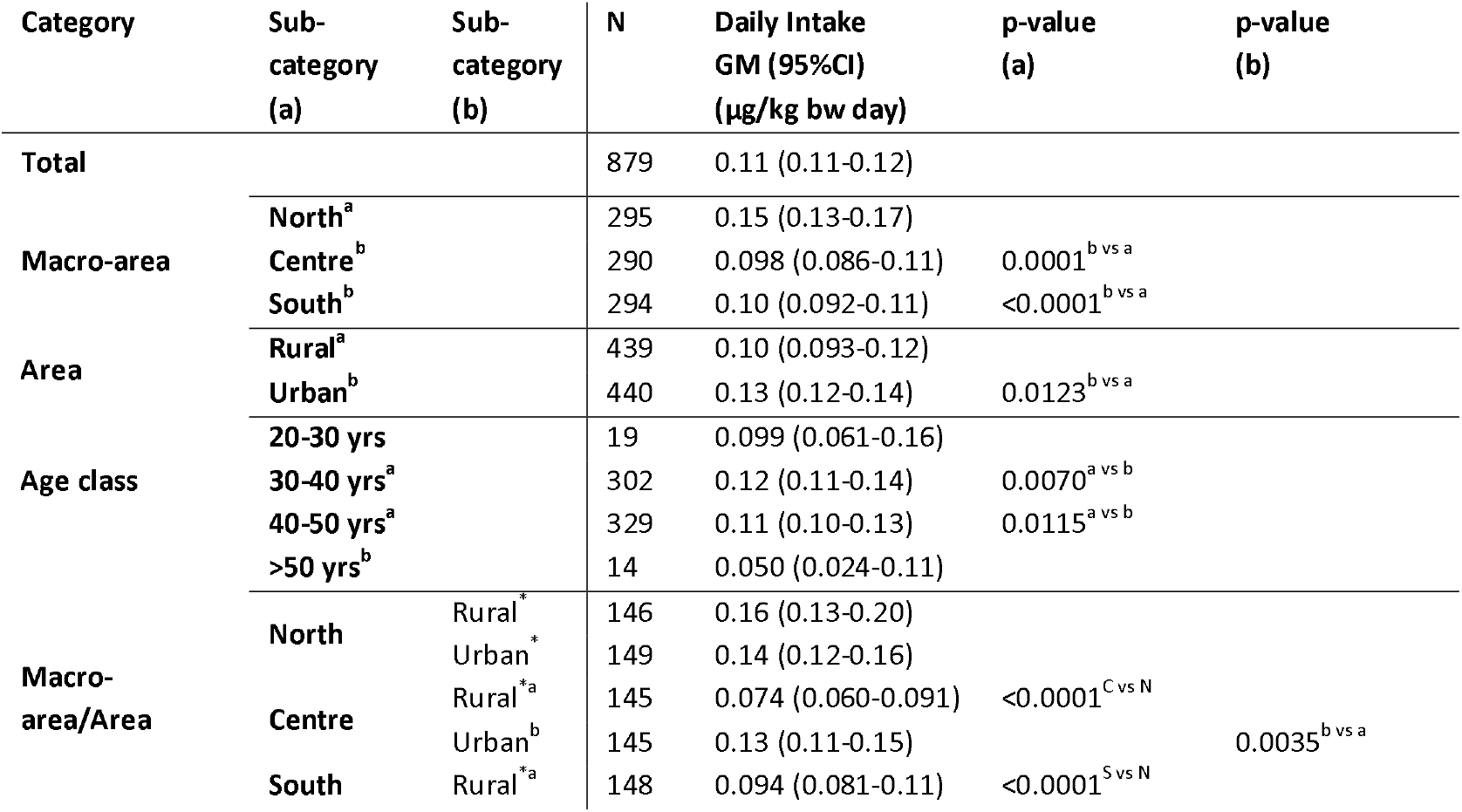
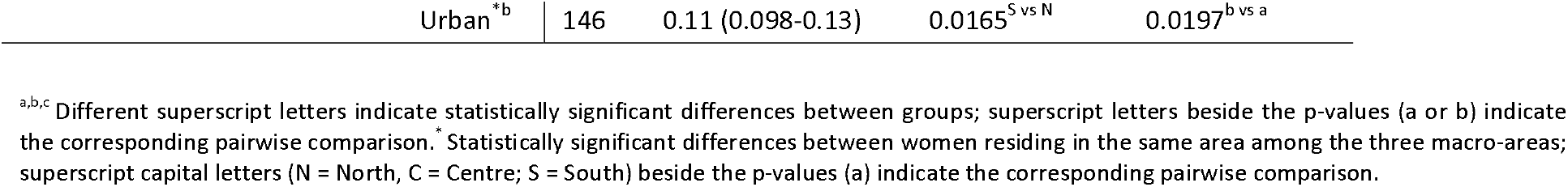
Estimated daily intake (µg/kg bw per day) of BPA in Italian women (N=879) according to residing areas and age.

The estimated total daily intake of BPA was significantly higher in the mothers living in Northern Italy and in urban areas, especially in Centre and South of Italy. In particular, women living in rural areas from Northern Italy had higher daily intake values than women in rural areas from Centre and South of Italy. Besides, urban Northern women had higher values than urban women from the South. In addition, women aged 30-50 years had higher daily intake values than women aged > 50 years.

### 3.4 Analysis of correlations

The sum of DEHP metabolites and BPA levels were positively correlated in the total population of Italian women, in Central Italy and in urban areas, both as unadjusted and adjusted values. Only for unadjusted values a positive correlation was found in South Italy and for adjusted values for women living in rural areas (**Table 10**).

**Table 10.** Correlations between the sum of DEHP metabolites and BPA levels as unadjusted and adjusted urinary concentrations in women (N=655) residing in different Italian macro-areas and areas. Rho coefficients and p-values are reported

|  | Unadjusted |  | Adjusted |  |
| --- | --- | --- | --- | --- |
|  | Rho | p-value | Rho | p-value |
| **TOT** | 0.1186 | **0.0004** | 0.1041 | **0.0018** |
| **North** | -0.0260 | 0.6537 | 0.0073 | 0.8998 |
| **Centre** | 0.2512 | **<0.0001** | 0.2561 | **<0.0001** |
| **South** | 0.2056 | **0.0004** | 0.1053 | 0.0700 |
| **Rural** | 0.0750 | 0.1139 | 0.1048 | **0.0269** |
| **Urban** | 0.1658 | **0.0004** | 0.1002 | **0.0344** |

Only for the sum of DEHP metabolite we found a positive correlation with age as adjusted levels, both in the total population and in Northern Italy although with very low Rho coefficient (**Supplementary Table 6**). A positive correlation was also observed in women from South Italy between adjusted sum of DEHP metabolite levels and BMI (**Supplementary Table 7**).

### 3.5 Reference values in women population

Reference values (RV95) in the total Italian women population were 101.40 and 30.59 µg/L for the sum of DEHP metabolites and BPA, respectively (**Table 11**). RV95 values for ΣDEHP were higher than the upper bound for the total population in South of Italy, especially the urban areas, and in younger mothers (age range 20-30 years). BPA RV95 values exceeded the upper bound for the general women population in the North of Italy and in rural areas, in particular the Northern ones.

**Table 11.** Reference values (RV95, P95(95% CI)) for the sum of DEHP metabolites and BPA in urine of women according to the residing area/macro-area and to age categories.

| Category | Sub-category (a) | Sub-category (b) | N | Σ DEHP RV95 [µg/L] (95%CI) | N | BPA RV95 [µg/L] (95%CI) |
| --- | --- | --- | --- | --- | --- | --- |
| Total |  |  | 892 | 101.40 (93.62-116.92) | 898 | 30.59 (25.46-37.53) |
| Macro-area | North |  | 300 | 96.42 (80.79-111.01) | 300 | 54.58 (38.63-66.76) |
|  | Centre |  | 295 | 94.33 (81.09-140.68) | 299 | 23.89 (19.67-31.54) |
|  | South |  | 297 | 123.71 (99.80-197.03) | 299 | 20.42 (16.64-25.18) |
| Area | Rural |  | 446 | 98.54 (87.16-139.03) | 448 | 39.41 (26.22-51.44) |
|  | Urban |  | 446 | 107.20 (94.72-125.30) | 450 | 27.16 (22.49-32.90) |
| Age class | 20-30 yrs |  | 19 | 187.58 (50.45-187.58) | 19 | 25.27 (13.37-25.27) |
|  | 30-40 yrs |  | 303 | 99.48 (85.06-122.78) | 304 | 30.44 (24.54-39.91) |
|  | 40-50 yrs |  | 333 | 100.73 (88.60-125.22) | 334 | 25.42 (21.74-38.73) |
|  | >50 yrs |  | 15 | 57.26 (45.58-57.26) | 15 | 15.05 (7.31-15.05) |
| Macro-area/Area | North | Rural | 150 | 96.23 (71.67-139.20) | 150 | 70.05 (51.04-94.05) |
|  |  | Urban | 150 | 97.94 (80.35-111.95) | 150 | 35.18 (24.52-51.97) |
|  | Centre | Rural | 148 | 94.86 (73.90-327.35) | 149 | 19.53 (13.38-26.41) |
|  |  | Urban | 147 | 94.51 (71.76-130.76) | 150 | 29.74 (21.04-49.37) |
|  | South | Rural | 148 | 113.37 (88.70-210.95) | 149 | 22.80 (15.84-38.62) |
|  |  | Urban | 149 | 151.59 (105.84-308.49) | 150 | 19.60 (14.81-26.22) |

## DISCUSSION

The LIFE PERSUADED project is a large HBM study that enrolled 900 mother-child pairs from 2015 to 2017 with the aim to establish the level of exposure to phthalates and BPA in the Italian population (La Rocca et al., 2018), where such data were still lacking.

The strength of the study is the first determination of the background and reference values in Italian women, useful in risk assessment (Vogel et al., 2019), and the evaluation of the co-exposure to DEHP and BPA. Moreover, the large number of mother-child pairs enrolled and the corresponding collected data during the LIFE PERSUADED project allow an accurate calculation of all these values also as a function of the different macro-areas and areas of residence.

DEHP metabolites were detected in the urine of 99% women and BPA in more than 95%, indicating that almost the entire Italian female adult population is exposed to these chemicals, in agreement with previous studies in other European countries (Cullen et al., 2017; Cutanda et al., 2015; Den Hond et al., 2015; Frederiksen et al., 2013; Kasper-Sonnenberg et al., 2012; Koppen et al., 2019; Larsson et al., 2014).

In Italian women, the geometric mean (GM) of ∑DEHP metabolites was similar to what observed in European women by Den Hond et al. in the DEMOCOPHES project (29.2 µg/L GM) (Den Hond et al., 2015); they showed that mothers from Romania had the highest concentration of DEHP (51.5 µg/L GM) while the lowest exposure was observed in United Kingdom (15.5 91 μg/L GM) (Den Hond et al., 2015). Hence, DEHP exposure in Italian mothers is within the range observed by European HBM studies in women. On the other hand, BPA exposure in Italian women was 5.79 μg/L, i.e., about 3 times higher than in other countries, as reported in a recent meta-analysis including 28,353 participants from different parts of the world with levels ranging from 0.81 to 3.50 μg/L (Colorado-Yohar et al., 2021). However, the concentration of BPA in Italian women here measured was similar to another study conducted in Italy in the Tuscany Chianti area involving 715 adults between 20 and 74 years old, where GM of BPA was 5.14 μg/L (Galloway et al., 2010), indicating a possible peculiarity of this territory. Moreover, since similar BPA levels were found in children enrolled in LIFE PERSUADED (Tait et al., 2021), it can be hypothesized that the Italian population is more exposed to BPA, as a possible consequence of lifestyles, eating habits or environmental contamination; in this regard an important contribution could come from the elaboration of data collected in LIFE PERSUADED questionnaires (unpublished data).

In this frame, we evaluated the impact of such internal levels on BPA daily intake to verify a possible exceedance of the total daily intake (TDI) of 4 ug/Kg bw per day established by EFSA in 2015 (EFSA Panel on Food Contact Materials, 2015). The daily intake estimation for DEHP was not calculated since the urinary concentration (as sum of metabolites) in Italian women was comparable to European data (Koppen et al., 2019) indicating TDI below the EFSA limit of 50 ug / kg bw per day. The results showed that the BPA daily intake in Italian women was 0.11 (0.11-0.12 GM, 95 CI) ug/kg bw per day, thus within the current TDI. However, due to collected evidence on adverse effects of BPA on the immune system, possibly representing the most sensitive target, EFSA is re-evaluating the TDI and proposing to reduce the value to 0.04 ng/kg bw per day (EFSA Panel on Food Contact Materials, 2021). In this scenario, all subjects enrolled in this HBM study have daily intake values well above the new proposed limit, i.e., 2750 and 4250 times higher for the mothers and their children, respectively. This observation raises concerns on BPA exposure in general, and in particular in Italy where internal levels are higher compared to other European and non-European countries, thus making urgent to adopt contrasting measures to limit exposure to this plasticizer.

With respect to geographical areas, DEHP exposure in women was higher in the Southern than in Central and Northern Italy with mothers’ exposure similar to that of their children (data previously reported in ref (Tait et al., 2020)). On the contrary, for BPA, different exposure patterns were observed in mothers and children (Tait et al., 2021); indeed, while mothers leaving in Northern Italy had the highest BPA exposure, children leaving in the South had the highest levels. Therefore, DEHP-like exposure in mother-child pairs based on area of residence appears to suggest common determinants of exposure independent of age and lifestyle; conversely, different determinants of exposure for young and adult populations may have resulted in diversified exposure to BPA.

No difference was observed in the levels of DEHP metabolites among women living in urban vs rural areas, similar to what reported for their children (Tait et al., 2020) and in other European countries (Cullen et al., 2017; Cutanda et al., 2015; Frederiksen et al., 2013; Larsson et al., 2014), with the exception of Ireland where MEOHP levels were found to be higher in women residing in the urban area (Cullen et al., 2017). BPA exposure was instead higher in Italian mothers living in urban areas with a trend similar to that observed in their children (Tait et al., 2021). This is probably due to different habits between differently urbanized areas that should be carefully considered in the evaluation of the risk.

The mothers that participated to this study had a median age of 41 years (38-44 interquartile range), they were relatively lean (median BMI of 22.27 kg/m2; 20.34-24.80 interquartile range) with a balanced distribution among macro-areas and urban vs rural areas. However, women living in Southern Italy and in rural areas were slightly younger and more overweight compared to mothers living in the North and Centre of Italy or in urban areas. Southern women showed a positive correlation between DEHP levels and BMI. In addition, a positive correlation was observed also between DEHP metabolites and age in the whole population and in the North of Italy, which is in contrast to what observed in children for which higher values were observed in the youngest (Tait et al., 2020).

The relative metabolic rates of DEHP metabolites in the Italian macro-areas showed that women living in Southern Italy had a more efficient metabolic conversion of MEHP into the secondary metabolites (MEHHP+MEOHP), with consequent lower percentages of molar MEHP, especially in women aged 30-50 yrs. Moreover, RMR2 was significantly different among women of the three macro-areas with a pattern Centre> North> South of Italy. Thus, in women both metabolic conversions differed across macro-areas whereas in children only RMR2 displayed differences but with a pattern North > Centre > South (Tait et al., 2020). Interestingly, a RMR1 pattern similar to women, decreasing from South to North, was observed in older children; hence, as previously hypothesized, RMR1 competence seems to increase with age while RMR2 mostly varies according to the residing geographic area (Tait et al., 2020). Although higher RMR2 was found in women aged 20-30 years, these data should be evaluated with caution because the number of subjects in the group is low; indeed, all other women > 30 years had comparable RMR2 values, thus supporting the previous hypothesis. Overall, the observed differences may be explained by considering that these metabolic conversions occur in the intestine and liver by enzymes of the cytochrome P450 chain, which have numerous polymorphisms within and between populations (Zanger and Schwab, 2013); probably this effect is even more evident in Italy due to the high genetic variability of the country (Capocasa et al., 2014).

RV95 values, which are useful parameters for the identification of the highest exposure scenario, were defined for both DEHP (∑MEHP, MEHHP, MEOHP) and BPA in Italian women aged from 24 to 67 years, equal to 101.40 µg/L and 30.59 µg/L, respectively. Compared to their children, RV95 for DEHP was higher in children (168 µg/L) whereas for BPA, children had a lower value compared to mothers (24.06 µg/L). Furthermore, the large sample size made it possible to derive RV95 values for subgroups of the female population based on geographical areas and age groups, making possible a more accurate assessment of exposure and risk.

Another interesting aspect of this study is the evaluation of co-exposure to DEHP and BPA in women; indeed, the concentrations of these plasticizers were positively correlated, in particular in Central and Southern Italy, suggesting possible common sources or an extensive use of plastics, resulting in a consequent cumulative exposure. This evidence is particularly relevant, considering the effects exerted by the mixture on the glucose metabolism, thyroid and cancer development exerted by the mixture and shown in in vivo studies (Baralic et al., 2021; Lin et al., 2011; Zhang et al., 2021; Zhang et al., 2022). In addition, the combined exposure of juvenile rats to DEHP and BPA, at same doses recorded in this HBM study, was associated to synergic action in the metabolic system and antagonism action in the reproductive and endocrine systems (Tassinari et al., 2021). In the adult population, the cumulative exposure has been associated with altered levels of thyroid hormones, which are particularly important for their role in energy metabolism and the reproductive system (Meeker and Ferguson, 2011; Park et al., 2017). Thus, the evidence gathered in the present HBM study further supports the concerns on possible adverse effects exerted by these plasticizers on young and adult populations, hopefully serving as a basis for policy regulators to introduce greater restrictions on their use.

## Conclusions

The data obtained from the LIFE PERSUADED project indicate ubiquitous exposure to DEHP and BPA for the Italian adult women. The results obtained in Italian mothers confirm the results in children, indicating women living in Southern Italy as the most exposed to phthalates and with less ability in processing the secondary metabolite MEHHP. Conversely, women living in Northern Italy and urban areas were more exposed to BPA. Overall, the area of residence is confirmed as an important determinant of exposure to plasticizers. Therefore, data on background levels, RV95 and dietary intake of DEHP and BPA in Italian mothers provide a contribution to the assessment of the risk associated to the exposure to plasticizers in adults in relation to the geographical area. Considering the health risk associated with exposure to plasticizers, these data should guide the implementation of policies aimed at reducing the public health impact of these chemicals and improving communication strategies to increase women’s awareness of protecting their health by reducing exposure to plasticizers.

## Data Availability

All data produced in the present work are contained in the manuscript

## Acknowledgments

This work was supported by the European Commission Directorate General Environment in the LIFE+ Programme, Environment Policy and Governance (grant number LIFE13 ENV/IT/000482).

## Components of the LIFE PERSUADED Project Group

**Istituto Superiore di Sanità, Rome, Italy:** Cinzia La Rocca, Luca Busani, Lucia Coppola, Gabriele Lori, Francesca Maranghi, Laura Narciso, Annalisa Silenzi, Sabrina Tait, Roberta Tassinari (Center for Gender-Specific Medicine); Francesca Romana Mancini, Roberta Urciuoli (Department of Infectious Diseases, Istituto Superiore di Sanità); Antonio Di Virgilio, Andrea Martinelli, Mauro Valeri (Centro Nazionale Sperimentale Benessere Animale, Istituto Superiore di Sanità, Rome, Italy)

**Istituto di Fisiologia Clinica, CNR, Pisa, Italy:** Amalia Gastaldelli, Graziano Barsotti, Emma Buzzigoli, Fabrizia Carli, Demetrio Ciociaro, Raffaele Conte, Veronica Della Latta, Graziella Distante, Melania Gaggini, Patrizia Landi, Anna Paola Pala, Chiara Saponaro.

**Dipartimento Pediatrico Universitario Ospedaliero “Bambino Gesu”, Rome, Italy:** Stefano Cianfarani, Barbara Baldini Ferroli, Giorgia Bottaro, Annalisa Deodati, Romana Marini, Giuseppe Scirè, Gian Luigi Spadoni

**Tor Vergata University, Rome, Italy;** Stefano Cianfarani, Daniela Germani

## Notes

### Competing Interest Statement

The authors have declared no competing interest.

### Clinical Protocols

https://www.mdpi.com/2218-1989/12/2/167

https://www.ncbi.nlm.nih.gov/pubmed/29974441

### Author Declarations

The whole study was approved by the Ethical Committee of the Istituto Superiore di Sanita'. The enrolled mothers signed an informed consent to provide a first-morning urine sample for the measurement of DEHP and BPA exposure and filled a questionnaire with information on residential environment, socio-demographic and lifestyle data as well as on food habits. Participants were assigned an alphanumeric code to guarantee anonymity (La Rocca et al. 2018).

## References

Baralic, K., Zivancevic, K., Jorgovanovic, D., Javorac, D., Radovanovic, J., Gojkovic, T., Buha Djordjevic, A., Curcic, M., Mandinic, Z., Bulat, Z., Antonijevic, B., Dukic-Cosic, D., 2021. Probiotic reduced the impact of phthalates and bisphenol A mixture on type 2 diabetes mellitus development: Merging bioinformatics with in vivo analysis. Food Chem Toxicol 154, 112325. doi: 10.1016/j.fct.2021.112325

Becker, K., Seiwert, M., Casteleyn, L., Joas, R., Joas, A., Biot, P., Aerts, D., Castano, A., Esteban, M., Angerer, J., Koch, H.M., Schoeters, G., Den Hond, E., Sepai, O., Exley, K., Knudsen, L.E., Horvat, M., Bloemen, L., Kolossa-Gehring, M., consortium, D., 2014. A systematic approach for designing a HBM pilot study for Europe. Int J Hyg Environ Health 217, 312–322. doi: 10.1016/j.ijheh.2013.07.004

Capocasa, M., Anagnostou, P., Bachis, V., Battaggia, C., Bertoncini, S., Biondi, G., Boattini, A., Boschi, I., Brisighelli, F., Calo, C.M., Carta, M., Coia, V., Corrias, L., Crivellaro, F., De Fanti, S., Dominici, V., Ferri, G., Francalacci, P., Franceschi, Z.A., Luiselli, D., Morelli, L., Paoli, G., Rickards, O., Robledo, R., Sanna, D., Sanna, E., Sarno, S., Sineo, L., Taglioli, L., Tagarelli, G., Tofanelli, S., Vona, G., Pettener, D., Destro Bisol, G., 2014. Linguistic, geographic and genetic isolation: a collaborative study of Italian populations. J Anthropol Sci 92, 201–231. doi: 10.4436/JASS.92001

Carli, F., Ciociaro, D., Gastaldelli, A., 2022. Assessment of Exposure to Di-(2-ethylhexyl) Phthalate (DEHP) Metabolites and Bisphenol A (BPA) and Its Importance for the Prevention of Cardiometabolic Diseases. Metabolites 12. doi: 10.3390/metabo12020167

Choi, J., Aarøe Mørck, T., Polcher, A., Knudsen, L.E., Joas, A., 2015. Review of the state of the art of human biomonitoring for chemical substances and its application to human exposure assessment for food safety. EFSA Supporting Publications 12. doi: 10.2903/sp.efsa.2015.EN-724

Chou, K., Wright, R.O., 2006. Phthalates in food and medical devices. J Med Toxicol 2, 126–135. doi: 10.1007/bf03161027

Colorado-Yohar, S.M., Castillo-Gonzalez, A.C., Sanchez-Meca, J., Rubio-Aparicio, M., Sanchez-Rodriguez, D., Salamanca-Fernandez, E., Ardanaz, E., Amiano, P., Fernandez, M.F., Mendiola, J., Navarro-Mateu, F., Chirlaque, M.D., 2021. Concentrations of bisphenol-A in adults from the general population: A systematic review and meta-analysis. Sci Total Environ 775, 145755. doi: 10.1016/j.scitotenv.2021.145755

Cullen, E., Evans, D., Griffin, C., Burke, P., Mannion, R., Burns, D., Flanagan, A., Kellegher, A., Schoeters, G., Govarts, E., Biot, P., Casteleyn, L., Castano, A., Kolossa-Gehring, M., Esteban, M., Schwedler, G., Koch, H.M., Angerer, J., Knudsen, L.E., Joas, R., Joas, A., Dumez, B., Sepai, O., Exley, K., Aerts, D., 2017. Urinary Phthalate Concentrations in Mothers and Their Children in Ireland: Results of the DEMOCOPHES Human Biomonitoring Study. Int J Environ Res Public Health 14. doi: 10.3390/ijerph14121456

Cutanda, F., Koch, H.M., Esteban, M., Sanchez, J., Angerer, J., Castano, A., 2015. Urinary levels of eight phthalate metabolites and bisphenol A in mother-child pairs from two Spanish locations. Int J Hyg Environ Health 218, 47–57. doi: 10.1016/j.ijheh.2014.07.005

Den Hond, E., Govarts, E., Willems, H., Smolders, R., Casteleyn, L., Kolossa-Gehring, M., Schwedler, G., Seiwert, M., Fiddicke, U., Castano, A., Esteban, M., Angerer, J., Koch, H.M., Schindler, B.K., Sepai, O., Exley, K., Bloemen, L., Horvat, M., Knudsen, L.E., Joas, A., Joas, R., Biot, P., Aerts, D., Koppen, G., Katsonouri, A., Hadjipanayis, A., Krskova, A., Maly, M., Morck, T.A., Rudnai, P., Kozepesy, S., Mulcahy, M., Mannion, R., Gutleb, A.C., Fischer, M.E., Ligocka, D., Jakubowski, M., Reis, M.F., Namorado, S., Gurzau, A.E., Lupsa, I.R., Halzlova, K., Jajcaj, M., Mazej, D., Tratnik, J.S., Lopez, A., Lopez, E., Berglund, M., Larsson, K., Lehmann, A., Crettaz, P., Schoeters, G., 2015. First steps toward harmonized human biomonitoring in Europe: demonstration project to perform human biomonitoring on a European scale. Environ Health Perspect 123, 255–263. doi: 10.1289/ehp.1408616

Duty, S.M., Ackerman, R.M., Calafat, A.M., Hauser, R., 2005. Personal care product use predicts urinary concentrations of some phthalate monoesters. Environ Health Perspect 113, 1530–1535. doi: 10.1289/ehp.8083

EFSA Panel on Food Contact Materials, E., Flavourings and Processing Aids, CEF, 2015. Scientific Opinion on the risks to public health related to the presence of bisphenol A (BPA) in foodstuffs. EFSA Journal 13. doi: 10.2903/j.efsa.2015.3978

EFSA Panel on Food Contact Materials, E., Processing, A., Silano, V., Barat Baviera, J.M., Bolognesi, C., Chesson, A., Cocconcelli, P.S., Crebelli, R., Gott, D.M., Grob, K., Lampi, E., Mortensen, A., Rivière, G., Steffensen, I.-L., Tlustos, C., Van Loveren, H., Vernis, L., Zorn, H., Cravedi, J.-P., Fortes, C., Tavares Poças, M.d.F., Waalkens-Berendsen, I., Wölfle, D., Arcella, D., Cascio, C., Castoldi, A.F., Volk, K., Castle, L., 2019. Update of the risk assessment of di-butylphthalate (DBP), butyl-benzyl-phthalate (BBP), bis(2-ethylhexyl)phthalate (DEHP), di-isononylphthalate (DINP) and di-isodecylphthalate (DIDP) for use in food contact materials. EFSA Journal 17, e05838. doi: https://doi.org/10.2903/j.efsa.2019.5838

EFSA Panel on Food Contact Materials, E.a.P.A., (CEP), 2021. Re-evaluation of the risks to public health related to the presence of bisphenol A (BPA) in foodstuffs.

Foulds, C.E., Trevino, L.S., York, B., Walker, C.L., 2017. Endocrine-disrupting chemicals and fatty liver disease. Nat Rev Endocrinol 13, 445–457. doi: 10.1038/nrendo.2017.42

Frederiksen, H., Nielsen, J.K., Morck, T.A., Hansen, P.W., Jensen, J.F., Nielsen, O., Andersson, A.M., Knudsen, L.E., 2013. Urinary excretion of phthalate metabolites, phenols and parabens in rural and urban Danish mother-child pairs. Int J Hyg Environ Health 216, 772–783. doi: 10.1016/j.ijheh.2013.02.006

Frederiksen, H., Skakkebaek, N.E., Andersson, A.M., 2007. Metabolism of phthalates in humans. Mol Nutr Food Res 51, 899–911. doi: 10.1002/mnfr.200600243

Galloway, T., Cipelli, R., Guralnik, J., Ferrucci, L., Bandinelli, S., Corsi, A.M., Money, C., McCormack, P., Melzer, D., 2010. Daily bisphenol A excretion and associations with sex hormone concentrations: results from the InCHIANTI adult population study. Environ Health Perspect 118, 1603–1608. doi: 10.1289/ehp.1002367

Guart, A., Bono-Blay, F., Borrell, A., Lacorte, S., 2011. Migration of plasticizers phthalates, bisphenol A and alkylphenols from plastic containers and evaluation of risk. Food Addit Contam Part A Chem Anal Control Expo Risk Assess 28, 676–685. doi: 10.1080/19440049.2011.555845

Halden, R.U., 2010. Plastics and health risks. Annu Rev Public Health 31, 179–194. doi: 10.1146/annurev.publhealth.012809.103714

Heindel, J.J., Vom Saal, F.S., Blumberg, B., Bovolin, P., Calamandrei, G., Ceresini, G., Cohn, B.A., Fabbri, E., Gioiosa, L., Kassotis, C., Legler, J., La Merrill, M., Rizzir, L., Machtinger, R., Mantovani, A., Mendez, M.A., Montanini, L., Molteni, L., Nagel, S.C., Parmigiani, S., Panzica, G., Paterlini, S., Pomatto, V., Ruzzin, J., Sartor, G., Schug, T.T., Street, M.E., Suvorov, A., Volpi, R., Zoeller, R.T., Palanza, P., 2015. Parma consensus statement on metabolic disruptors. Environ Health 14, 54. doi: 10.1186/s12940-015-0042-7

Ho, K.L., Yuen, K.K., Yau, M.S., Murphy, M.B., Wan, Y., Fong, B.M., Tam, S., Giesy, J.P., Leung, K.S., Lam, M.H., 2017. Glucuronide and Sulfate Conjugates of Bisphenol A: Chemical Synthesis and Correlation Between Their Urinary Levels and Plasma Bisphenol A Content in Voluntary Human Donors. Arch Environ Contam Toxicol 73, 410–420. doi: 10.1007/s00244-017-0438-1

Huang, C.C., Yang, C.Y., Su, C.C., Fang, K.M., Yen, C.C., Lin, C.T., Liu, J.M., Lee, K.I., Chen, Y.W., Liu, S.H., Huang, C.F., 2021. 4-Methyl-2,4-bis(4-hydroxyphenyl)pent-1-ene, a Major Active Metabolite of Bisphenol A, Triggers Pancreatic beta-Cell Death via a JNK/AMPKalpha Activation-Regulated Endoplasmic Reticulum Stress-Mediated Apoptotic Pathway. Int J Mol Sci 22. doi: 10.3390/ijms22094379

Kasper-Sonnenberg, M., Koch, H.M., Wittsiepe, J., Wilhelm, M., 2012. Levels of phthalate metabolites in urine among mother-child-pairs-results from the Duisburg birth cohort study, Germany. Int J Hyg Environ Health 215, 373–382. doi: 10.1016/j.ijheh.2011.09.004

Koch, H.M., Bolt, H.M., Angerer, J., 2004. Di(2-ethylhexyl)phthalate (DEHP) metabolites in human urine and serum after a single oral dose of deuterium-labelled DEHP. Arch Toxicol 78, 123–130. doi: 10.1007/s00204-003-0522-3

Koppen, G., Govarts, E., Vanermen, G., Voorspoels, S., Govindan, M., Dewolf, M.C., Den Hond, E., Biot, P., Casteleyn, L., Kolossa-Gehring, M., Schwedler, G., Angerer, J., Koch, H.M., Schindler, B.K., Castano, A., Lopez, M.E., Sepai, O., Exley, K., Bloemen, L., Knudsen, L.E., Joas, R., Joas, A., Schoeters, G., Covaci, A., 2019. Mothers and children are related, even in exposure to chemicals present in common consumer products. Environ Res 175, 297–307. doi: 10.1016/j.envres.2019.05.023

Kumar, M., Sarma, D.K., Shubham, S., Kumawat, M., Verma, V., Prakash, A., Tiwari, R., 2020. Environmental Endocrine-Disrupting Chemical Exposure: Role in Non-Communicable Diseases. Front Public Health 8, 553850. doi: 10.3389/fpubh.2020.553850

La Rocca, C., Maranghi, F., Tait, S., Tassinari, R., Baldi, F., Bottaro, G., Buzzigoli, E., Carli, F., Cianfarani, S., Conte, R., Deodati, A., Gastaldelli, A., Pala, A.P., Raffaelli, A., Saponaro, C., Scire, G., Spadoni, G.L., Busani, L., Group, L.P.P., 2018. The LIFE PERSUADED project approach on phthalates and bisphenol A biomonitoring in Italian mother-child pairs linking exposure and juvenile diseases. Environ Sci Pollut Res Int 25, 25618–25625. doi: 10.1007/s11356-018-2660-4

Larsson, K., Ljung Bjorklund, K., Palm, B., Wennberg, M., Kaj, L., Lindh, C.H., Jonsson, B.A., Berglund, M., 2014. Exposure determinants of phthalates, parabens, bisphenol A and triclosan in Swedish mothers and their children. Environ Int 73, 323–333. doi: 10.1016/j.envint.2014.08.014

Lin, Y., Wei, J., Li, Y., Chen, J., Zhou, Z., Song, L., Wei, Z., Lv, Z., Chen, X., Xia, W., Xu, S., 2011. Developmental exposure to di(2-ethylhexyl) phthalate impairs endocrine pancreas and leads to long-term adverse effects on glucose homeostasis in the rat. Am J Physiol Endocrinol Metab 301, E527–538. doi: 10.1152/ajpendo.00233.2011

Lobstein, T., Brownell, K.D., 2021. Endocrine-disrupting chemicals and obesity risk: A review of recommendations for obesity prevention policies. Obes Rev 22, e13332. doi: 10.1111/obr.13332

Meeker, J.D., Ferguson, K.K., 2011. Relationship between urinary phthalate and bisphenol A concentrations and serum thyroid measures in U.S. adults and adolescents from the National Health and Nutrition Examination Survey (NHANES) 2007-2008. Environ Health Perspect 119, 1396–1402. doi: 10.1289/ehp.1103582

Mengozzi, A., Carli, F., Biancalana, E., Della Latta, V., Seghieri, M., Gastaldelli, A., Solini, A., 2019. Phthalates Exposure as Determinant of Albuminuria in Subjects With Type 2 Diabetes: A Cross-Sectional Study. J Clin Endocrinol Metab 104, 1491–1499. doi: 10.1210/jc.2018-01797

Park, C., Choi, W., Hwang, M., Lee, Y., Kim, S., Yu, S., Lee, I., Paek, D., Choi, K., 2017. Associations between urinary phthalate metabolites and bisphenol A levels, and serum thyroid hormones among the Korean adult population - Korean National Environmental Health Survey (KoNEHS) 2012-2014. Sci Total Environ 584-585, 950-957. doi: 10.1016/j.scitotenv.2017.01.144

Tait, S., Carli, F., Busani, L., Buzzigoli, E., Della Latta, V., Deodati, A., Fabbrizi, E., Gaggini, M., Maranghi, F., Tassinari, R., Toffol, G., Cianfarani, S., Gastaldelli, A., La Rocca, C., Group, L.P.P., 2020. Biomonitoring of Bis(2-ethylhexyl)phthalate (DEHP) in Italian children and adolescents: Data from LIFE PERSUADED project. Environ Res 185, 109428. doi: 10.1016/j.envres.2020.109428

Tait, S., Carli, F., Busani, L., Ciociaro, D., Della Latta, V., Deodati, A., Fabbrizi, E., Pala, A.P., Maranghi, F., Tassinari, R., Toffol, G., Cianfarani, S., Gastaldelli, A., La Rocca, C., Life Persuaded Project, G., 2021. Italian Children Exposure to Bisphenol A: Biomonitoring Data from the LIFE PERSUADED Project. Int J Environ Res Public Health 18. doi: 10.3390/ijerph182211846

Tassinari, R., Tait, S., Busani, L., Martinelli, A., Valeri, M., Gastaldelli, A., Deodati, A., La Rocca, C., Maranghi, F., The Life Persuaded Project Group, 2021. Toxicological Assessment of Oral Co-Exposure to Bisphenol A (BPA) and Bis(2-ethylhexyl) Phthalate (DEHP) in Juvenile Rats at Environmentally Relevant Dose Levels: Evaluation of the Synergic, Additive or Antagonistic Effects. Int J Environ Res Public Health 18, 4584. doi: 10.3390/ijerph18094584

Vogel, N., Conrad, A., Apel, P., Rucic, E., Kolossa-Gehring, M., 2019. Human biomonitoring reference values: Differences and similarities between approaches for identifying unusually high exposure of pollutants in humans. Int J Hyg Environ Health 222, 30–33. doi: 10.1016/j.ijheh.2018.08.002

WHO, 1996. Biological monitoring of chemical exposure in the workplace, Guidelines. World Health Organization, Geneva.

Wittassek, M., Angerer, J., 2008. Phthalates: metabolism and exposure. Int J Androl 31, 131–138. doi: 10.1111/j.1365-2605.2007.00837.x

Yilmaz, B., Terekeci, H., Sandal, S., Kelestimur, F., 2020. Endocrine disrupting chemicals: exposure, effects on human health, mechanism of action, models for testing and strategies for prevention. Rev Endocr Metab Disord 21, 127–147. doi: 10.1007/s11154-019-09521-z

Yoshihara, S., Mizutare, T., Makishima, M., Suzuki, N., Fujimoto, N., Igarashi, K., Ohta, S., 2004. Potent estrogenic metabolites of bisphenol A and bisphenol B formed by rat liver S9 fraction: their structures and estrogenic potency. Toxicol Sci 78, 50–59. doi: 10.1093/toxsci/kfh047

Zanger, U.M., Schwab, M., 2013. Cytochrome P450 enzymes in drug metabolism: regulation of gene expression, enzyme activities, and impact of genetic variation. Pharmacol Ther 138, 103–141. doi: 10.1016/j.pharmthera.2012.12.007

Zhang, X., Cheng, C., Zhang, G., Xiao, M., Li, L., Wu, S., Lu, X., 2021. Co-exposure to BPA and DEHP enhances susceptibility of mammary tumors via up-regulating Esr1/HDAC6 pathway in female rats. Ecotoxicol Environ Saf 221, 112453. doi: 10.1016/j.ecoenv.2021.112453

Zhang, X., Guo, N., Jin, H., Liu, R., Zhang, Z., Cheng, C., Fan, Z., Zhang, G., Xiao, M., Wu, S., Zhao, Y., Lu, X., 2022. Bisphenol A drives di(2-ethylhexyl) phthalate promoting thyroid tumorigenesis via regulating HDAC6/PTEN and c-MYC signaling. J Hazard Mater 425, 127911. doi: 10.1016/j.jhazmat.2021.127911

