## Supplementary for "Women exposure to Di(2-ethylhexyl)phthalate (DEHP) and bisphenol A (BPA) from different residing areas in Italy: data from the LIFE PERSUADED project"

Appendix

**Supplementary Table 1** - DEHP metabolites level in urine samples of mothers residing in the North (N=300), Centre (N=299) or South (N=299) of Italy. In the table are reported the geometric mean (GM) with the 95% CI, the median (P50) and the IQ range (P25-P75). Both urinary (µg/L) and creatinine-adjusted concentrations (µg/g) are shown.

| Metabolite | Macro-area | Unit | GM (95% CI) | P50 (P25-P75) | p-value |
| --- | --- | --- | --- | --- | --- |
| MEHP | **North^a^** | µg/L | 4.40 (4.04-4.81) | 4.82 (3.14-6.74) | 0.0009^a vs b^ |
|  | **Centre^a^** |  | 4.73 (4.32-5.19) | 4.93 (3.39-7.47) | 0.0392^a vs b^ |
|  | **South^b^** |  | 4.81 (4.30-5.37) | 5.58 (3.90-7.96) |  |
|  | **North** | µg/g crea | 4.30 (3.91-4.73) | 4.41 (2.83-6.58) |  |
|  | **Centre** |  | 4.33 (3.88-4.82) | 4.16 (2.79-6.79) |  |
|  | **South** |  | 4.29 (3.79-4.85) | 4.84 (3.15-7.31) |  |
| MEHHP | **North^a^** | µg/L | 16.18 (14.69-17.82) | 16.28 (9.37-29.52) | <0.0001^a vs b^ |
|  | **Centre^a^** |  | 16.82 (15.06-18.79) | 17.56 (10.05-29.32) | 0.0008^a vs b^ |
|  | **South^b^** |  | 21.50 (19.36-23.87) | 21.69 (12.51-38.57) |  |
|  | **North^a^** | µg/g crea | 15.59 (14.31-16.98) | 15.18 (10.08-24.97) | 0.0019^a vs b^ |
|  | **Centre^a^** |  | 15.28 (13.90-16.79) | 15.14 (9.62-23.21) | 0.0005^a vs b^ |
|  | **South^b^** |  | 19.03 (17.32-20.91) | 18.00 (11.73-30.20) |  |
| MEOHP | **North^a^** | µg/L | 4.95 (4.51-5.44) | 4.99 (2.92-8.50) | <0.0001^a vs b^ |
|  | **Centre^a^** |  | 4.81 (4.31-5.36) | 5.13 (2.73-8.64) | <0.0001^a vs b^ |
|  | **South^b^** |  | 7.09 (6.39-7.87) | 6.98 (4.29-12.05) |  |
|  | **North^a^** | µg/g crea | 4.79 (4.41-5.20) | 4.69 (3.10-6.92) | 0.0448^a vs b^ |
|  | **Centre^b^** |  | 4.39 (4.00-4.81) | 4.31 (2.70-6.87) | <0.0001^b vs c^ |
|  | **South^c^** |  | 6.33 (5.75-6.96) | 6.02 (4.11-9.38) | <0.0001^c vs a^ |

^a,b,c^ Different superscript letters indicate statistically significant differences between groups; superscript letters beside the p-values indicate the corresponding pairwise comparison.

**Supplementary Table 2** - DEHP metabolites level in urine samples of mothers residing in rural (N=448) or in urban (N=450) Italian areas. In the table are reported the geometric mean (GM) with the 95% CI, the median (P50) and the IQ range (P25-P75). Both urinary (µg/L) and creatinine-adjusted concentrations (µg/g) are shown.

| Metabolite | Area | Unit | GM (95% CI) | P50 (P25-P75) |
| --- | --- | --- | --- | --- |
| MEHP | **Rural^a^** | µg/L | 4.74 (4.38-5.14) | 5.12 (3.52-7.54) |
|  | **Urban^b^** |  | 4.54 (4.20-4.92) | 5.04 (3.22-7.21) |
|  | **Rural** | µg/g crea | 4.28 (3.91-4.68) | 4.49 (2.90-7.20) |
|  | **Urban** |  | 4.33 (3.97-4.73) | 4.50 (2.96-6.52) |
| MEHHP | **Rural** | µg/L | 17.77 (16.31-19.35) | 17.89 (10.47-31.46) |
|  | **Urban** |  | 18.27 (16.77-19.91) | 18.08 (10.49-31.69) |
|  | **Rural** | µg/g crea | 15.89 (14.73-17.14) | 15.56 (10.17-25.01) |
|  | **Urban** |  | 17.24 (16.02-18.56) | 16.47 (10.49-27.68) |
| MEOHP | **Rural** | µg/L | 5.48 (5.04-5.96) | 5.54 (3.25-9.31) |
|  | **Urban** |  | 5.57 (5.12-6.08) | 5.66 (3.14-10.06) |
|  | **Rural^a^** | µg/g crea | 4.93 (4.58-5.30) | 4.81 (3.05-7.60) |
|  | **Urban^b^** |  | 5.29 (4.90-5.71) | 5.17 (3.22-8.32) |

^a,b,c^ Different superscript letters indicate statistically significant differences between groups; superscript letters beside the p-values indicate the corresponding pairwise comparison.

**Supplementary Table 3** - DEHP metabolites level in urine samples of mothers (N=898) residing in rural or in urban areas in the three macro-areas. In the table are reported the geometric mean (GM) with the 95% CI, the median (P50) and the IQ range (P25-P75). Both urinary (µg/L) and creatinine-adjusted concentrations (µg/g) are shown.

| Macro-Area (MA) | Metabolite | Area | Unit | GM (95% CI) | P50 (P25-P75) | p-value (Area) | p-value (MA) |
| --- | --- | --- | --- | --- | --- | --- | --- |
| North | **MEHP** | **Rural^*^** | µg/L | 7.54 (6.79-8.37) | 4.69 (2.92-6.21) |  |  |
|  |  | **Urban** |  | 7.20 (6.57-7.90) | 5.00 (3.30-7.21) |  |  |
|  |  | **Rural^*^** | µg/g crea | 7.13 (6.31-8.05) | 4.16 (2.56-6.41) |  |  |
|  |  | **Urban** |  | 7.17 (6.50-7.90) | 4.52 (3.10-6.72) |  |  |
|  | **MEHHP** | **Rural^*^** | µg/L | 15.30 (13.08-17.90) | 16.43 (9.60-29.84) |  |  |
|  |  | **Urban^*^** |  | 15.34 (13.16-17.89) | 14.88 (8.50-28.26) |  |  |
|  |  | **Rural^*^** | µg/g crea | 14.46 (12.55-16.67) | 15.65 (9.46-25.13) |  |  |
|  |  | **Urban^*^** |  | 15.27 (13.45-17.34) | 14.85 (10.15-24.97) |  |  |
|  | **MEOHP** | **Rural^*^** | µg/L | 4.81 (4.17-5.55) | 4.93 (2.97-8.34) |  |  |
|  |  | **Urban^*^** |  | 5.00 (4.39-5.69) | 5.05 (2.89-8.94) |  |  |
|  |  | **Rural^*^** | µg/g crea | 4.54 (3.99-5.16) | 4.71 (3.01-6.69) |  | 0.0310 **^N vs C^** |
|  |  | **Urban^*^** |  | 4.97 (4.48-5.52) | 4.48 (3.25-7.00) |  |  |
| Centre | **MEHP** | **Rural^*^** | µg/L | 8.17 (7.35-9.09) | 4.87 (3.56-7.83) |  |  |
|  |  | **Urban** |  | 7.15 (6.47-7.90) | 4.93 (3.15-7.23) |  |  |
|  |  | **Rural^*^** | µg/g crea | 7.23 (6.35-8.23) | 4.03 (2.76-7.23) |  |  |
|  |  | **Urban** |  | 6.87 (6.10-7.74) | 4.42 (3.04-6.30) |  |  |
|  | **MEHHP** | **Rural^*^** | µg/L | 15.26 (12.62-18.46) | 15.93 (9.81-29.61) |  |  |
|  |  | **Urban^*^** |  | 17.12 (14.85-19.75) | 18.58 (10.93-29.28) |  |  |
|  |  | **Rural^a*^** | µg/g crea | 13.29 (11.25-15.70) | 13.90 (8.74-21.76) |  |  |
|  |  | **Urban^b^** |  | 16.33 (14.61-18.24) | 16.51 (10.38-24.99) |  |  |
|  | **MEOHP** | **Rural^*^** | µg/L | 4.74 (4.06-5.53) | 4.87 (2.90-8.19) |  |  |
|  |  | **Urban^*^** |  | 4.66 (3.88-5.59) | 5.48 (2.65-8.92) |  |  |
|  |  | **Rural^*^** | µg/g crea | 4.14 (3.64-4.70) | 3.86 (2.62-5.51) |  |  |
|  |  | **Urban^*^** |  | 4.44 (3.82-5.16) | 4.68 (2.74-7.73) |  |  |
| South | **MEHP** | **Rural^a*^** | µg/L | 8.77 (7.70-10.00) | 5.87 (4.25-8.38) |  | <0.0001 **^S vs N^**  0.0102 **^S vs C^** |
|  |  | **Urban^b^** |  | 7.36 (6.24-8.70) | 5.27 (3.11-7.21) | 0.0157^b vs a^ |  |
|  |  | **Rural^a*^** | µg/g crea | 7.74 (6.70-8.93) | 5.04 (3.73-8.13) |  | 0.0022 **^S vs N^**  0.0154 **^S vs C^** |
|  |  | **Urban^b^** |  | 6.89 (5.80-8.19) | 4.49 (2.63-6.39) | 0.0301^b vs a^ |  |
|  | **MEHHP** | **Rural^*^** | µg/L | 21.00 (18.43-23.93) | 21.50 (13.47-35.83) |  | 0.0043 **^S vs N^**  0.0050 **^S vs C^** |
|  |  | **Urban^*^** |  | 20.15 (16.66-24.37) | 22.55 (11.59-42.91) |  | 0.0018 **^S vs N^**  0.0261 **^S vs C^** |
|  |  | **Rural^*^** | µg/g crea | 18.30 (16.35-20.48) | 18.19 (12.99-28.32) |  | 0.0192 **^S vs N^**  0.0009 **^S vs C^** |
|  |  | **Urban^*^** |  | 18.32 (15.42-21.77) | 17.55 (10.79-33.11) |  | 0.0214 **^S vs N^** |
|  | **MEOHP** | **Rural^*^** | µg/L | 7.23 (6.34-8.24) | 7.31 (4.35-10.92) |  | <0.0001 **^S vs N^**  <0.0001 **^S vs C^** |
|  |  | **Urban^*^** |  | 7.45 (6.44-8.63) | 6.63 (4.25-12.36) |  | 0.0004 **^S vs N^**  0.0004 **^S vs C^** |
|  |  | **Rural^*^** | µg/g crea | 6.34 (5.67-7.08) | 5.90 (4.24-9.30) |  | 0.0001 **^S vs N^**  <0.0001 **^S vs C^** |
|  |  | **Urban** |  | 6.82 (5.98-7.78) | 6.06 (4.01-9.70) |  | 0.0011 **^S vs N^**  0.0002 **^S vs C^** |

^a,b,c^ Different superscript letters indicate statistically significant differences between areas; superscript letters beside the p-values (Area) indicate the corresponding pairwise comparison.

^*^ Statistically significant differences between women residing in the same area among the three macro-areas; superscript capital letters (N = North, C = Centre; S = South) beside the p-values (by MA) indicate the corresponding pairwise comparison.

**Supplementary Table 4** - Relative metabolic rates and percentage fractions of DEHP metabolites in women according to age categories. Data are expressed as medians (IQ range).

| Age |  | P50 (P25-P75) | p-value |
| --- | --- | --- | --- |
| 20-30 (N=19) | RMR1 | 4.05 (1.74-7.28) |  |
|  | RMR2^a^ | 3.99 (3.35-4.85) |  |
|  | %MEHP | 19.81 (12.08-36.54) |  |
|  | %MEHHP^a^ | 55.84 (47.72-65.55) |  |
|  | %MEOHP^a^ | 21.51 (18.53-34.14) |  |
| 30-40 (N=303) | RMR1^a^ | 4.94 (3.03-7.78) | 0.0164^a vs b^ |
|  | RMR2^b^ | 3.24 (2.35-4.35) | 0.0238^b vs a^ |
|  | %MEHP^a^ | 16.83 (11.40-24.84) | 0.0164^a vs b^ |
|  | %MEHHP | 61.95 (54.21-70.24) |  |
|  | %MEOHP^b^ | 19.24 (15.47-24.31) | 0.0047^b vs a^ |
| 40-50 (N=333) | RMR1^a^ | 4.96 (3.20-7.67) | 0.0117^a vs b^ |
|  | RMR2^b^ | 3.10 (2.29-4.11) | 0.0107^b vs a^ |
|  | %MEHP^a^ | 16.78 (11.53-23.83) | 0.0117^a vs b^ |
|  | %MEHHP^b^ | 62.65 (54.63-70.06) | 0.0460^b vs a^ |
|  | %MEOHP^b^ | 19.46 (15.12-23.55) | 0.0022^b vs a^ |
| >50 (N=15) | RMR1^b^ | 3.70 (1.81-5.13) |  |
|  | RMR2^b^ | 2.87 (2.42-3.75) | 0.0218^b vs a^ |
|  | %MEHP^b^ | 21.27 (16.32-35.62) |  |
|  | %MEHHP | 58.27 (46.56-65.95) |  |
|  | %MEOHP^b^ | 18.05 (13.84-18.96) | 0.0017^c vs a^ |

^a,b,c^ Different superscript letters indicate statistically significant differences among groups; superscript letters beside the p-values (area) indicate the corresponding pairwise comparison.

**Supplementary Table 5** - Relative metabolic rates and percentage fractions of DEHP metabolites in women according to BMI categories. Data are expressed as medians (IQ range).

| BMI |  | P50 (P25-P75) | p-value |
| --- | --- | --- | --- |
| <25 (N=672) | RMR1 | 4.87 (2.86-7.44) |  |
|  | RMR2^a^ | 3.14 (2.28-4.19) |  |
|  | %MEHP | 17.04 (11.84-25.93) |  |
|  | %MEHHP | 62.33 (52.58-70.02) |  |
|  | %MEOHP^a^ | 19.25 (14.87-23.50) | 0.0054 ^a vs b^ |
| 25-30 (N=150) | RMR1 | 5.05 (3.41-7.92) |  |
|  | RMR2 | 3.15 (2.38-4.36) |  |
|  | %MEHP | 16.53 (11.21-22.68) |  |
|  | %MEHHP^a^ | 63.97 (55.36-69.89) |  |
|  | %MEOHP^a^ | 19.82 (15.89-23.35) | 0.0364 ^a vs b^ |
| >30 (N=56) | RMR1 | 4.69 (3.21-7.97) |  |
|  | RMR2^b^ | 3.34 (2.59-4.85) | 0.0266 ^b vs a^ |
|  | %MEHP | 17.58 (11.15-23.77) |  |
|  | %MEHHP^b^ | 58.94 (53.86-64.27) | 0.0423 ^b vs a^ |
|  | %MEOHP^b^ | 20.86 (17.46-27.05) |  |

^a,b,c^ Different superscript letters indicate statistically significant differences among groups; superscript letters beside the p-values (area) indicate the corresponding pairwise comparison.

**Supplementary Table 6** – Correlation between the sum of DEHP metabolites or BPA levels as adjusted urinary concentration and age of enrolled women (N=655) in Italian macro-areas and areas. Rho coefficients and p-values are reported

|  | N | Σ DEHP metabolites | | BPA | |
| --- | --- | --- | --- | --- | --- |
|  | N | **Rho** | **p-value** | **Rho** | **p-value** |
| TOT | 655 | 0.0795 | **0.0422** | 0.0753 | 0.0541 |
| North | 254 | 0.1358 | **0.0304** | 0.0286 | 0.6496 |
| Centre | 193 | 0.0901 | 0.2102 | 0.0895 | 0.2157 |
| South | 208 | 0.0527 | 0.4497 | 0.1086 | 0.1186 |
| Rural | 319 | 0.0439 | 0.4347 | 0.0519 | 0.3558 |
| Urban | 336 | 0.1029 | 0.0604 | 0.0450 | 0.4110 |

**Supplementary Table 7** – Correlation of the sum of DEHP metabolites and BPA as adjusted urinary concentrations with BMI in women (N=879).

|  | N | Σ DEHP metabolites | | BPA | |
| --- | --- | --- | --- | --- | --- |
|  |  | **Rho** | **p-value** | **Rho** | **p-value** |
| TOT | 879 | 0.0217 | 0.5218 | -0.0469 | 0.1653 |
| North | 295 | -0.0136 | 0.8159 | 0.0544 | 0.3516 |
| Centre | 290 | -0.0808 | 0.1729 | -0.0825 | 0.1611 |
| South | 294 | 0.1180 | **0.0442** | -0.0409 | 0.4860 |
| Rural | 439 | 0.0401 | 0.4028 | -0.0575 | 0.2290 |
| Urban | 440 | 0.0063 | 0.8961 | -0.0200 | 0.6753 |
